# How Rare is Rare? *TNFAIP3* Variants and the High Collective Burden of Haploinsufficiency

**DOI:** 10.64898/2026.04.20.26350987

**Authors:** Danica M. Lee, Urekha Karri, Charlotte V. Cuff, Casey A. Rimland, Priyamvada Guha Roy, Kader Cetin Gedik, Josh Owens, Manuel Carpio Tumba, Sam Chiang, Prabal Chhibbar, Hey Chong, Michael Ero, Kyr Goyette, Eric Hanson, Akuti Kethri, Kyle Jones, Younglang Lee, Erika Owsley, Connie Ng, Liva Pfuhler, Camille Sambar, Li Yang, Jiebiao Wang, Guido H. Falduto, Pui Lee, Jeffrey A. Sparks, Jishnu Das, Yiming Luo, Daniella M. Schwartz

**Author notes:** Contributed equally, co-corresponding authors (Jishnu Das, Yiming Luo, Daniella M. Schwartz). Corresponding Authors: Daniella M. Schwartz University of Pittsburgh, Departments of Medicine and Immunology 200 Lothrop St. Pittsburgh, PA 15213 Jishnu Das University of Pittsburgh, Department of Immunology, Center for Systems Immunology Yiming Luo Department of Medicine, Columbia University Medical Center.

## Abstract

**Background:** Monogenic diseases are considered rare, yet many remain underdiagnosed when clinical manifestations are heterogeneous. A20 haploinsufficiency (HA20) is an early-onset inborn error of immunity (IEI) caused by heterozygous *TNFAIP3* variants that dysregulate inflammatory signaling, producing diverse immune phenotypes.

**Methods:** We analyzed variants in all human haploinsufficiency (HI) disease genes in gnomAD, All of Us (AoU), and UK Biobank (UKBB), applying refined loss-of-function predictors. Rare *TNFAIP3* variants (allele frequency <0.01%) were classified as predicted loss-of-function (pLOF) or functionally validated highly predicted pathogenic missense (fHPPM). Clinical associations were tested through meta-phenome-wide association studies (PheWAS) and phenome-set enrichment analysis (PSEA) and, for *TNFAIP3*, validated in a University of Pittsburgh referral cohort and a single center biobank at Mass General Brigham (MGB).

**Findings:** Deleterious variants in human HI disease genes, including IEI-HI genes, occurred frequently despite strong constraint and were significantly associated with immune disease risk. *TNFAIP3* pLOF variant prevalences were as high as ∼1:24,000; combined pLOF + fHPPM prevalences reached ∼1:9,000. PheWAS and PSEA linked rare *TNFAIP3* variants to diverse immune phenotypes. In a referral cohort (18 patients, 9 families), missense variants conferred hypomorphism with intermediate immunophenotypes. In the MGB biobank, 29 individuals carried hypomorphic *TNFAIP3* variants without a HA20 diagnosis; nearly two-thirds had phenotypes consistent with or plausibly related to A20 hypomorphism, suggesting penetrance exceeds 50%. Only 4 had received HA20-directed immunomodulatory therapy.

**Interpretation:** Deleterious IEI-HI variants are far more common than reported and are associated with immune dysregulation; hypomorphic *TNFAIP3* variants are common and highly penetrant with variable expressivity. Genotype-first analysis of institutional biobanks can identify untreated IEI-HI patients. These findings establish proof-of-concept that haploinsufficiency diseases are pervasively underrecognized. Patients with early-onset or treatment-refractory immune disease should be prioritized for genetic testing; biobank callback amendments may expedite diagnosis, monitoring, and precision therapy.

## Introduction

Monogenic conditions are often assumed to be rare. Recent work suggests that many could be underdiagnosed and their collective population prevalence underestimated, particularly for disorders with heterogeneous clinical phenotypes^1^. However, prior studies have focused on incompletely penetrant alleles with well-defined phenotypes and have lacked definitive validation in independent clinical cohorts^2,3^. The relevance of these findings to heterogeneous immune conditions and the links between population-wide variant burden and clinical disease burden remain unclear. One such monogenic disease, A20 haploinsufficiency (HA20), is an early□onset inflammatory syndrome caused by high□penetrance, heterozygous germline variants in *TNFAIP3*^4^*. TNFAIP3* encodes the multifunctional ubiquitin-editing enzyme A20^5,6^. HA20 patients exhibit dysregulation of TNF-NF-κB, inflammasome, and interferon signaling pathways and present with diverse phenotypes including autoinflammation, autoimmunity, lymphoproliferation, and recurrent infection^7^.

Since its discovery, HA20 has been considered ultra-rare with fewer than 200 reported cases^7,8^, most involving frameshift, truncating, deletion, or splice variants. Missense variants are reported to cause HA20 but are challenging to validate in the absence of clinically standardized assays, leading to potential under-ascertainment. Moreover, genetic sequencing is not universally accessible, and older patients are often diagnosed only through family□based segregation testing^8^. Together, these factors introduce substantial ascertainment bias and lead to missed opportunities for intervention.

We hypothesized that HA20 prevalence is substantially underestimated in the U.S. and globally, and that rigorous validation through functional, clinical, and population-scale analyses could have broad implications for haploinsufficient immune diseases. We quantified the prevalence and clinical impact of deleterious variants predicted to cause haploinsufficiency disease at U.S. and global population levels. Focusing on *TNFAIP3* variants across three population resources, we applied rare variant collapsing phenome-wide association studies (PheWAS) to uncover strong associations with immune disease. We validated these findings through functional characterization of variants, deep immunophenotyping of a University of Pittsburgh referral cohort, and analysis of the Mass General Brigham (MGB) hospital-based biobank, providing rigorous, multimodal evidence that a heterogeneous immune disease is underrecognized and establishing proof-of-concept for underrecognition of haploinsufficiency diseases at the population level.

## Methods

(*Detailed Methods available in **Supp. Appendix***)

### Participants and Data Sources

Whole genome sequencing and electronic health record (EHR)-linked data were analyzed from the All of Us Program version 8 (AoU v8; n=414,830), UK Biobank (UKBB; n=425,966), and gnomAD v4.0 (n=807,162) ^9–11^. AoU and UKBB participants were not selected for specific diseases; gnomAD attempts to exclude participants with severe Mendelian disease^9–11^. Diagnosis codes were mapped to phecodes (Phecode v1.2) for phenome-wide association studies (PheWAS). Genetic ancestry was inferred using principal component analysis (**Supp. Fig. 1A**); ancestry categories were defined by each source database and used as provided without additional reclassification^12–14^.

### University of Pittsburgh HA20 Cohort

Individuals with suspected pathogenic *TNFAIP3* variants (pLOF and missense) were referred for evaluation under IRB-approved STUDY22060106; all participants provided informed consent. Variant classification followed ACMG criteria and incorporated NF-κB luciferase assays (see below)^15^. Immunologic profiling included S100A8/9, S100A12, IL-18 (ELISA), type I interferon gene score (NanoString), IL-1β (ELISA), and basal A20 expression (Western)^16–19^.

### Mass General Biobank Cohort

The MGBB is a prospective institutional EHR-linked biorepository (MGB IRB #2019P000264) that includes data from 67,576 participants who underwent whole exome/genome sequencing. *TNFAIP3* variant carriers were then identified, then variants were classified as described below. EHR of participants carrying pLOF, HPPM, or Pitt cohort missense variants were then reviewed for phenotypes consistent with HA20 spectrum disease or plausibly related to A20 hypomorphism.

### Variant Classification (population databases)

For analysis of all haploinsufficiency genes, variants were defined based on predicted loss-of-function (pLoF): frameshift, nonsense, or splice with a SpliceAI > 0.9 and/or damaging missense (CADD-Phred > 30). For *TNFAIP3*-specific analysis, variants with minor allele frequency (MAF) <0.01% were classified as: pLOF or highly predicted pathogenic missense (HPPM; CADD-Phred≥30, SIFT=deleterious, PolyPhen=probably damaging, no homozygosity) (**Supp. Fig. 1B**) ^20–22^. Functional validation used NF-κB luciferase assays (see below). U.S. and global counts were extrapolated from 2026 population estimates.

### Phenome-wide Association and Comparative Clinical Phenotype Analysis

PheWAS for pLOF and HPPM carriers were performed separately in each biobank under the REGENIE framework using logistic regression with Firth correction^23^. Models were adjusted for sex, age, and the first 10 genetic ancestry principal components, with the leave-one-chromosome-out (LOCO) predictor from whole-genome regression included as an offset. Fixed-effects meta-analysis combining AoU and UKBB results was then performed using REMETA^24^. Phenome-wide statistical significance was defined using a Bonferroni-corrected threshold of p < 2.75 x 10^-5^ (0.05/1817; phecodes with ≥20 cases). Enrichment of inflammatory phenotypes was assessed by applying gene set enrichment analysis (GSEA) to phecodes ranked by their association Z scores. HPPM and pLOF carriers (AoU) were sex□ and age□matched with virtual controls (3:1 control:case, ‘MatchIt’, R) lacking rare *TNFAIP3* variants. Targeted AoU EHR review incorporated diagnosis dates and code provenance to refine phenotypes.

### NF-κB luciferase assay

A20□ deficient HEK293 cells were transiently transfected with NF□κB reporter plasmids (Cignal) and GFP□tagged A20, then stimulated with TNFα (20 ng/mL). After 5 hours, luciferase activity was measured, with firefly luciferase normalized to renilla luciferase (RLU) and expressed as fold induction versus wild□type A20.

### Basal A20 expression

Peripheral blood mononuclear cells (PBMCs) lysates were tested for basal A20 expression (Western blot) via immunoblotting with A20 N-terminus antibody (CST), visualization by chemiluminescence, and normalization to β-actin.

### Statistical Analysis

Analyses were performed in R and GraphPad Prism. Genetic constraint metrics were calculated as previously described^25,26^. Prevalence estimates used descriptive statistics with 95% confidence intervals (Prism). Odds ratios used Fisher’s exact test (R). Group comparisons and incidence rate ratios (IRRs) used negative binomial regression (R). Biomarker comparisons used Kruskal-Wallis with Dunn’s correction or Brown-Forsythe and Welch ANOVA depending on results of normality testing (Prism).

## Results

### Predicted deleterious variants in haploinsufficiency genes across population databases

Haploinsufficiency (HI) diseases like CTLA4, SOCS1, and A20 haploinsufficiency are considered ultra-rare (<1:1,000,000) based on reported cases^7,27–29^. However, the collective prevalence of HI disease variants in population databases has not been calculated. We modified stringent LOF (loss-of-function) predictors to identify high-confidence deleterious variants across non-HI genes, HI genes, and immune-related HI (IHI) genes in gnomAD (**Supp. Data 1**). Although IHI genes showed the greatest mutational constraint, they harbored more predicted deleterious variants than expected from reported case numbers (**Fig. 1A-B**). This included predicted LOF (pLOF: nonsense, frameshift, splice null) and predicted damaging missense (CADD-Phred > 30) variants.

**Figure 1.**
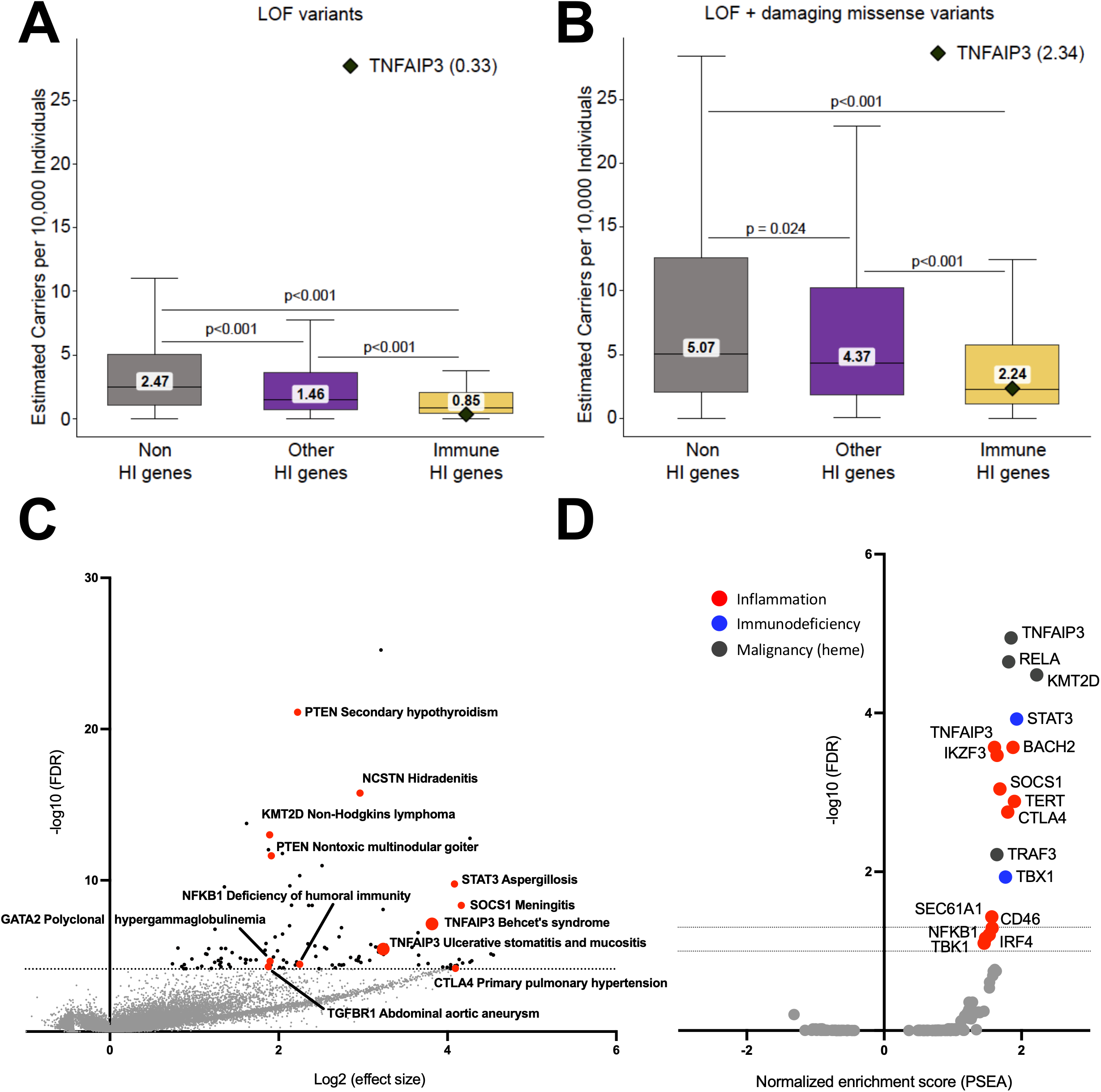
**A,B.** Box plots show prevalence (number of individuals carrying a single variant per 10,000 individuals, gnomAD v4) with loss-of-function, LOF (A) and damaging missense (B) variants. Non-haploinsufficiency (HI) genes, grey; non-immune HI disease-causal genes, purple; inborn error of immunity HI disease-causal genes, gold. LOF variants defined as truncating (nonsense), frameshift, splice null (Splice AI > 0.9). Damaging missense defined as CADD-Phred > 30. Per-person prevalence was calculated by dividing the heterozygous allele count by total number of individuals callable for the gene. (p-values, Mann-Whitney with Bonferroni correction). Black diamond, *TNFAIP3*. **C.** Volcano plot shows meta-PheWAS (AoU, UKBB) for individuals with pLOF variants in 48 immune-related haploinsufficiency (IHI) genes with reported inborn error of immunity phenotypes (IEI). Each data point represents a specific gene-phecode association. Black data points represent significant IEI-HI associations, gray data points represent nonsignificant associations. Red data points are labeled IEI-HI genes of interest. Gray dotted line, false discovery rate (FDR)-adjusted threshold for statistical significance (p<0.05). **D.** Volcano plot depicts phenotype set enrichment analysis (PSEA) results for 48 IEI-HI genes, using a curated set of phecodes categorized as inflammatory, immunodeficient, or hematologic malignancy (“phenotype cassettes”). Gray dotted lines, stringent and permissive FDR-adjusted thresholds for significance (p<0.05, p<0.1).

### Haploinsufficiency-IEI genes broadly confer significant population-scale disease burden

The mismatch between the frequency of predicted-damaging variants and reported IHI cases suggests incomplete penetrance, subclinical variable expressivity, or significant underdiagnosis of IHI diseases. To systematically test the clinical impact of IHI variants, we conducted an unbiased rare-variant meta-phenome-wide association study (PheWAS) using a collapsing framework that pooled variant carriers in AoU and UKBB, given that most variants were seen in 1-2 individuals. To maintain stringency, we restricted our analysis to pLOF (nonsense, frameshift, splice null) variants in 48 gold-standard IHI genes with established IEI phenotypes (IEI-HI; **Supp. Data 2-3**)^30^. Two-thirds (32/48, 66.7%) showed significant phenotypic associations (FDR < 0.05), and over half (25/48, 52.1%) were consistent with expected IEI phenotypes or mechanistically plausible (**Fig. 1C**). Even under stringent Bonferroni correction, half of all IEI-HI genes (24/48, 50%) retained significant associations, and 41.7% (20/48) remained consistent or plausible. Beyond recapitulating established phenotypes, our analysis identified unexpected gene-phenotype associations, including TERT with glomerulonephritis and TLR3 with chronic venous hypertension.

To test for net effect of all 48 IEI-HI genes on immune dysregulation at population-scale, we categorized 1,876 phecodes as inflammatory, immunodeficient, or hematologic malignancy (**Supp. Data 4**). We compared LOF carriers versus non-carriers using phenotype set enrichment analysis (PSEA), providing an estimate of inflammatory/immune disease burden related to IEI-HI genes (**Fig. 1D; Supp. Data 5**). One-third of genes (16/48) significantly increased the pooled risk of inflammatory disease (11/48), immunodeficient (2/48), or hematologic malignancy (4/48), with *TNFAIP3* in two categories. Together, these data demonstrate that LOF mutations in haploinsufficiency-IEI genes confer extremely high risk of human immunologic disease, including potential new phenotypes for future investigation.

### Inactivating TNFAIP3 variants are abundant in US and global populations

*TNFAIP3* is an IEI-HI gene with 100% reported penetrance for LOF variants^7^ but fewer than 200 reported cases, despite a higher-than-expected prevalence of damaging variants (**Fig. 1A-B**) and strong PheWAS and PSEA associations at a population level (**Fig. 1C-D**). We therefore focused on *TNFAIP3* as an exemplar of IEI-HI variants, first validating pLOF variant prevalence findings in All of Us (AoU) and UK Biobank (UKBB) (**Table 1, Supp. Fig. 1B**). The estimated prevalence was approximately 1:25,927 in the U.S. (95% CI, 1:15,960–1:42,119) and 1:23,740 globally (95% CI, 1:16,989–1:33,172). Extrapolated to 2026 populations, this corresponds to ∼13,461 individuals in the United States and ∼349,620 worldwide. All pLOF alleles failed to suppress TNF-induced NF-κB activation relative to wild-type A20, consistent with haploinsufficiency (**Fig. 2A; Supp. Data 7,8**). pLOF variants from gnomAD, AoU, and UKBB were numerically enriched in the zinc finger 7 (ZnF7) domain relative to other domains (**Supp. Fig. 1C)**, consistent with ZnF7’s critical role in restraining inflammatory signaling^31,32^.

**Figure 2.**
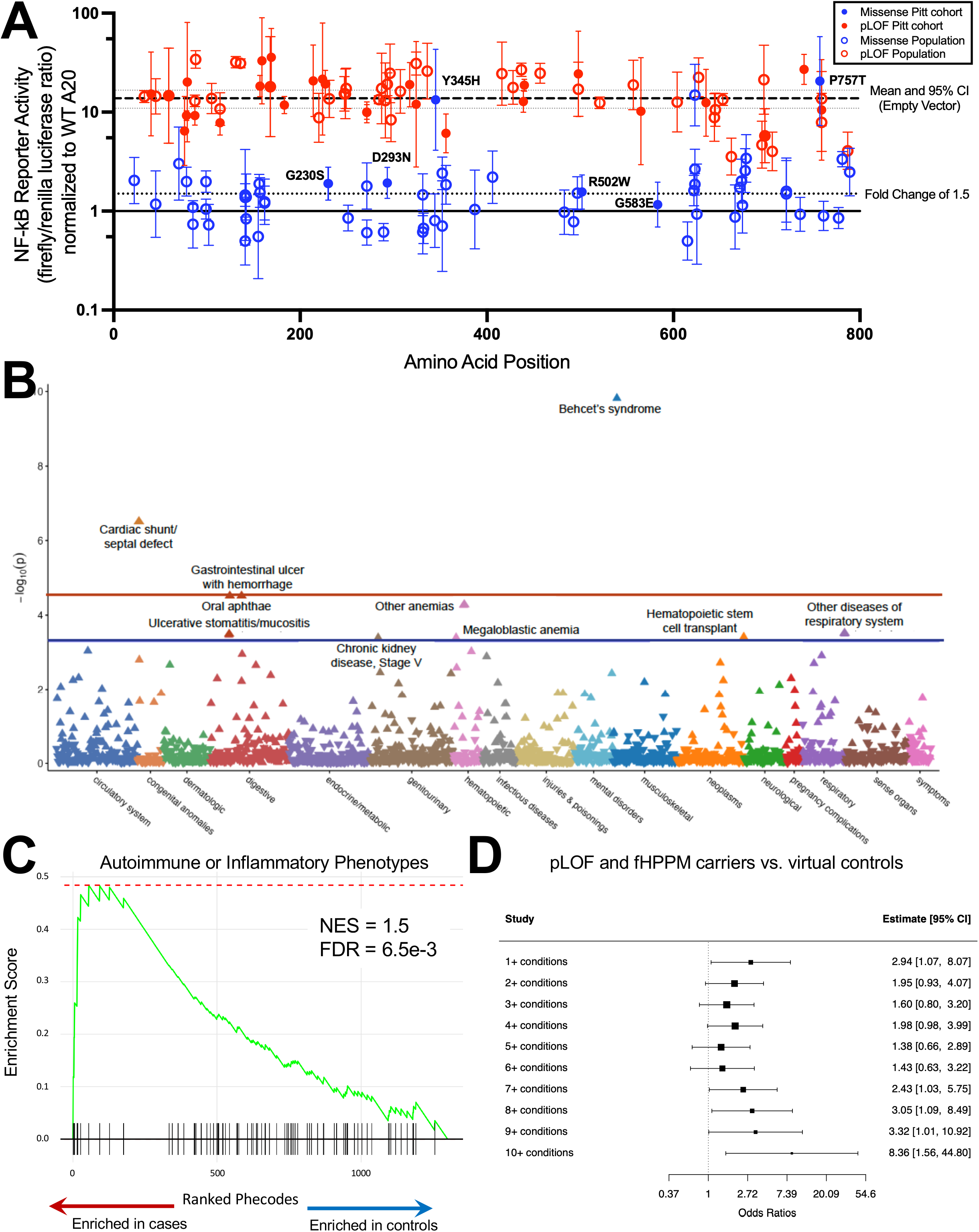
**A.** A20-deficient HEK293 cells were co-transfected with NF-κB luciferase reporter plasmids and mutant or WT A20 plasmid or EV, then stimulated with TNFα for 5 hours. Scatterplot shows geometric mean of NF-κB reporter activity ± geometric SD firefly/renilla luciferase RLU for pLOF and HPPM *TNFAIP3* variants, normalized to WT A20 (solid line). Red open circle, pLOF variants from population databases; blue open circle, HPPM variants from population databases; red solid circle, pLOF variants from Pittsburgh cohort; blue solid circle, ACMG pathogenic missense variants from Pittsburgh cohort. Dashed line, mean RLU for EV; dotted lines, 95% CI for EV. **B.** Manhattan plot shows meta-PheWAS (AoU, UKBB) for individuals with predicted loss-of-function (pLOF) and functionally validated highly predicted pathogenic missense (HPPM) *TNFAIP3* variants. Each data point represents a phecode; color delineates its phecode category. Red line, Bonferroni-corrected threshold for statistical significance (p<2.75×10□□); blue, suggestive threshold (p<5×10□^4^). **C.** PSEA plot shows enrichment of immune/inflammatory phecodes (NES = 1.5, FDR-adjusted p = 0.0065) in AoU and UKBB carriers of pLOF and functionally validated HPPM *TNFAIP3* variants relative to other individuals. **D.** Forest plot shows odds ratio of having ≥1 HA20-related diagnosis (Fisher’s exact test) at each cumulative threshold in individuals from AoU with linked EHR data and pLOF and functionally validated HPPM *TNFAIP3* variants (n=43) vs. sex- and age-matched controls (n=129). Black square, each threshold effect size estimate; size of square, weight based on precision; whiskers, 95% CI for each estimate. HEK, human epithelial kidney; SD, standard deviation; EV, empty vector; RLU, relative light unit; PheWAS, phenome-wide association study, pLOF, predicted loss-of-function; HPPM, highly predicted pathogenic missense; AoU, All of Us; UKBB, United Kingdom Biobank; Phecode, phenotype code; PSEA, phenotype set enrichment analysis; NES, normalized enrichment score.

**Table 1.**
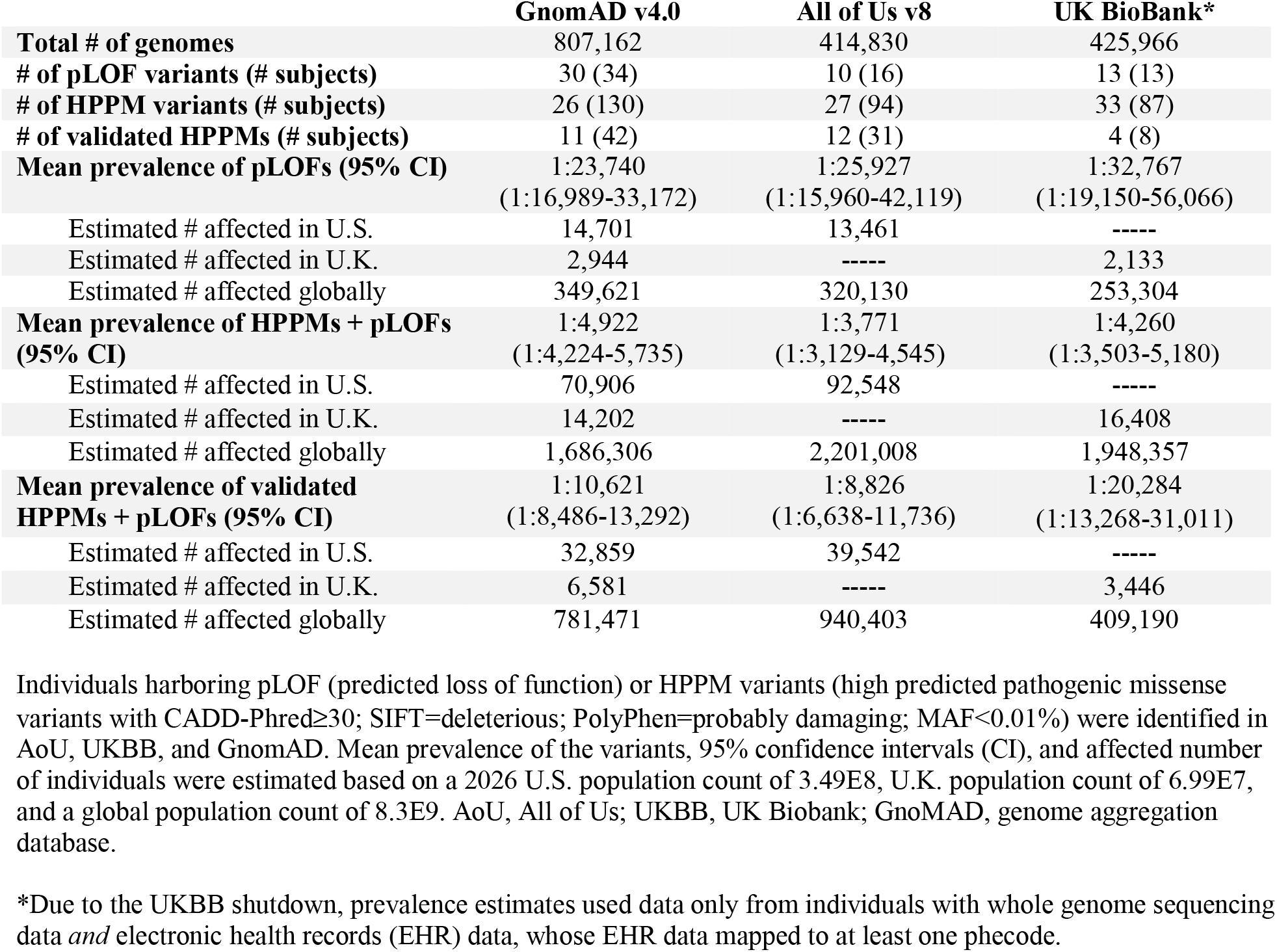
Prevalence estimates of pLOF or HPPM *TNFAIP3* variants in the US (AoU), UK (UKBB), and globally (GnomAD)

### Hypomorphic missense TNFAIP3 variants are prevalent

To avoid underestimating the burden of *TNFAIP3*-related disease by excluding pathogenic missense variants, we next incorporated a stringent class of “highly predicted pathogenic missense” (HPPM) variants defined by multi-tool concordance (CADD ≥30, SIFT=deleterious, PolyPhen=probably damaging), rarity (MAF<0.01%), and absence of homozygosity. Across AoU v8, UKBB, and gnomAD v4.0, we identified 43 unique HPPM variants (AoU 27; UKBB 33; gnomAD 26). Of these, 1 HPPM variant (p.Phe623Ser) failed to suppress TNF-induced NF-κB activation. The other 42 HPPM alleles retained the ability to suppress TNF-induced NF-κB activation, but 18 variants (42.9%) were hypomorphic, exhibiting ∼1.5–3-fold less suppression than wild-type A20 (**Fig. 2A**). HPPM variants were significantly depleted in nonfunctional linker regions and enriched in the ZnF4 domain, which mediates K48-linked ubiquitination and degradation of proinflammatory signaling proteins (**Supp. Fig. 1D**)^33^. When functionally validated HPPM and pLOF carriers were combined, we observed 47 potentially affected individuals in AoU v8, 76 in gnomAD v4, and 21 in UKBB (**Table 1**), corresponding to a U.S. prevalence of approximately 1:8,826 (95% CI, 1:6,638–1:11,736; AoU v8) and a global prevalence of approximately 1:10,621 (95% CI, 1:8,486–1:13,292; gnomAD), corresponding to about 39,542 individuals in the United States and 781,471 worldwide.

### Rare, deleterious TNFAIP3 variants are highly penetrant

Focusing on all hypomorphic variants (LOF and functionally validated HPPM), we next asked whether these alleles are associated with significant disease burden at a population level. We pooled carriers of all functionally deleterious *TNFAIP3* variants (pLOF and HPPM) in AoU and UKBB for meta-PheWAS as we had previously done for pLOF variants alone (**Fig. 1C, 2B, Supp. Fig. 2A; Supp. Data 6**). Collapsed variant carrier status was strongly associated with hallmark HA20 phenotypes, including Behçet’s disease (adjusted odds ratio [aOR] 23466.7; 95% CI, 987-557936), gastrointestinal ulcers with hemorrhage (aOR, 227.1; 95% CI, 13.6-3796.6), and oral aphthae (aOR, 188.8; 95% CI, 19.3-1841.2), surpassing a Bonferroni-corrected significance threshold (p<2.75×10□□). Additional immune diagnoses reported in HA20 were also enriched, e.g., megaloblastic anemia (aOR, 58927.3; 95% CI, 540.3-6426390) and ulcerative mucositis (aOR, 406; 95% CI, 19.5-8433.7)^34^. When analyzed separately, meta-PheWAS showed multiple associations (aOR181.1-103086) surpassing stringent significance for LOF variants while showing weaker but still HA20-relevant associations (aOR 9.9-99.5) for hypomorphic missense variants, consistent with a gradient of disease risk across variant classes (**Supp. Fig. 2A-B**).

To test for broader immune associations without prespecifying phenotypes, we categorized 1,876 phecodes as inflammatory versus non-inflammatory (**Supp. Data 4**) and observed significant enrichment of inflammatory phecodes (normalized enrichment score [NES] = 1.5, FDR = 0.0065) (**Fig. 2C**). Recognizing potential cross-resource heterogeneity in EHR curation, we complemented PheWAS with clinician-informed phenotyping in AoU v8: among 47 variant (pLOF and functionally validated HPPM, fHPPM) carriers, 43 had EHR data, which we compared with 3:1 age- and sex-matched controls (n=129). Variant carriers had a 52% higher incidence rate of potential HA20-related diagnoses (**Supp. Data 9**, incidence rate ratio: 1.52, 95% CI [1.09, 2.14], p=0.014) and higher odds of having >1 such diagnosis (**Fig. 2D; Supp. Fig. 2C**). Across these orthogonal approaches, hypomorphic *TNFAIP3* variants markedly increase the risk of potential immune/inflammatory disease.

### Missense TNFAIP3 variants in 18 HA20 patients from 9 unrelated families

Limitations of population-scale data include lack of familial segregation and primary samples for deep immunophenotyping, as well as inconsistency of EHR data. Considering these limitations and the risk-gradient for hypomorphic missense vs. LOF variants, we investigated a referral cohort to probe the functional and clinical consequences of A20 hypomorphism. Eighteen patients from nine unrelated families with inflammatory phenotypes and suspected hypomorphic *TNFAIP3* missense variants underwent clinician-led phenotyping, family-based segregation, and functional testing; variants were classified as pathogenic or likely pathogenic per American College of Medical Genetics (ACMG) criteria (**Supp. Table 1, Supp. Appendix**).

### Clinical, functional, and immunological consequences of HA20-causal missense variants

All *TNFAIP3* variants were described in relation to NM_001270508.2. Two variants (c.1033T>C, p.Tyr345His; c.2269C>A, p.Pro757Thr) were strongly hypomorphic for NF-κB suppression, comparable to nonsense/frameshift alleles; three (c.688G>A, p.Gly230Ser; c.1504C>T, p.Arg502Trp; c.877G>A, p.Asp293Asn) were weakly hypomorphic, and one (c.1748G>A, p.Gly583Glu) suppressed NF-κB similarly to wild type A20 (**Fig. 2A**). Basal A20 protein was reduced in PBMCs from c.2269C>A (p.Pro757Thr) and c.1748G>A (p.Gly583Glu) carriers, indicating expression-level hypomorphism even when the NF-κB readout was preserved in overexpression-based assays (**Fig. 3A**). Patients with deleterious missense variants showed “intermediate” activation of A20-regulated immune pathways: higher than healthy volunteers but lower than those with pLOF variants (**Fig. 3B-F**). These included biomarkers of myeloid activation (S100A8/9, S100A12), inflammasome activation (IL-18, LPS-induced IL-1β production), interferon signaling (type I interferon gene score)^16,18,19^. Compared with pLOF carriers from the same referral cohort (n = 48), missense carriers had lower frequencies of genital ulcers, autoantibodies, and immune cytopenias; there was a trend towards higher frequencies of lung involvement, central nervous system disease, sicca symptoms, and immunodeficiency phenotypes (**Supp. Table 2)**. Together, these data support a spectrum of A20 hypomorphism among missense variants, ranging from functional to expression-level defects, and accompanied by a distinguishable but partially overlapping immune dysregulatory endophenotype, consistent with the risk gradient seen at the population level.

**Figure 3.**
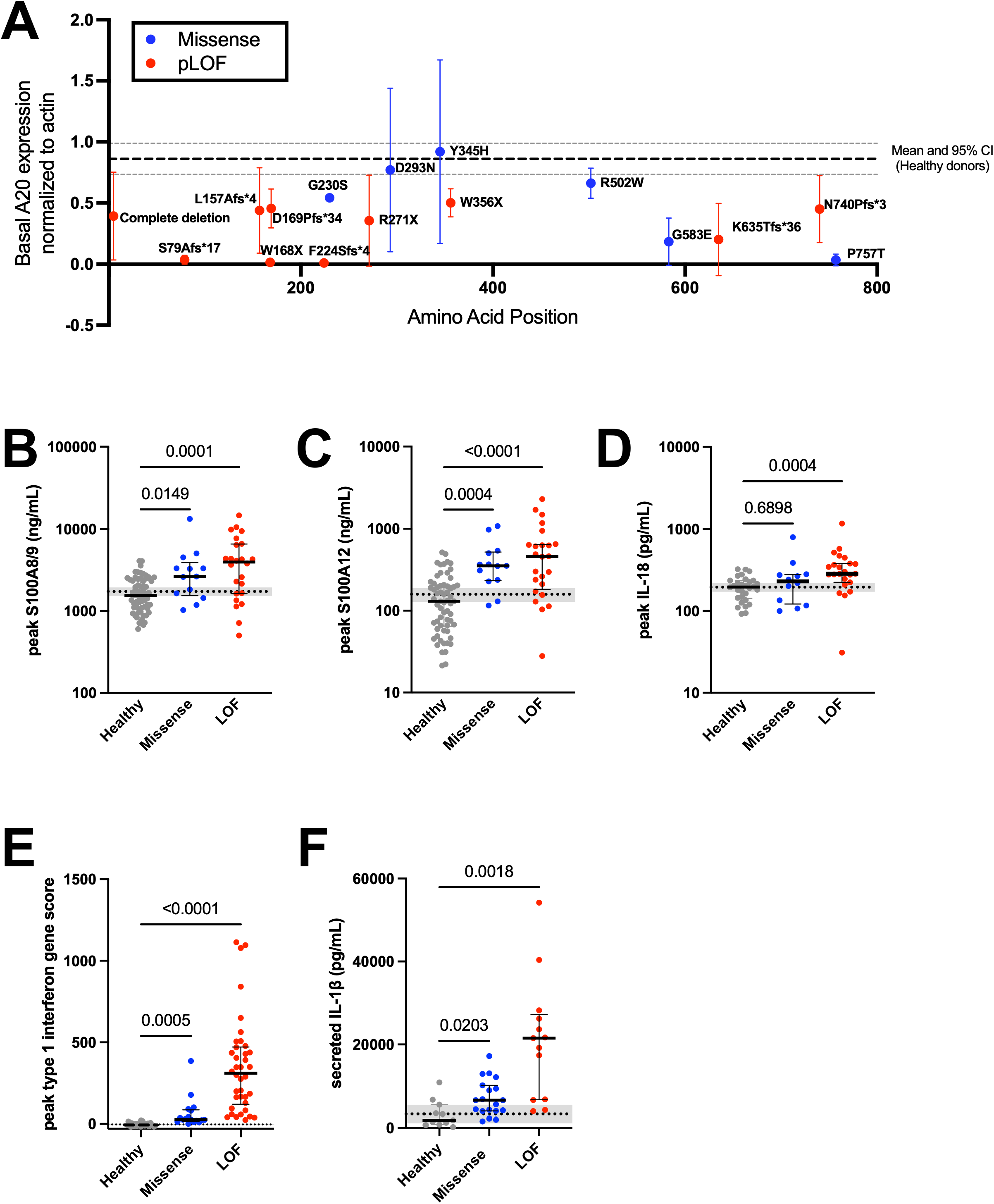
**A.** Scatterplot shows mean ± SD basal A20 expression (Western blot, normalized to β-actin) in PBMCs from patients with nonsense/frameshift (red solid circle) and pathogenic missense (blue solid circle) variants in the Pittsburgh cohort. Dashed line, mean basal A20 expression in healthy volunteers; dotted lines, 95% CI in healthy volunteers. **B-E.** Plasma S100A8/9 levels (B, Kruskal-Wallis p<0.0001), plasma S100A12 levels (C, Kruskal-Wallis p<0.0001), plasma IL-18 levels (D, Kruskal-Wallis p=0.0009), whole blood type I interferon stimulated gene scores (E, Kruskal-Wallis p<0.0001) from HA20 patients and healthy volunteers. **F.** PBMCs from HA20 patients and healthy volunteers were stimulated with LPS for four hours before IL-1β concentration in supernatant was measured via ELISA (Brown-Forsythe p=0.0004; Welch’s ANOVA p=0.0005). Healthy volunteers, grey solid circle; patients with LOF (nonsense/frameshift/null) variants, red solid circle; patients with missense variants, blue solid circle; p-values, Kruskal-Wallis test with Dunn’s multiple correction. Black line, median with interquartile range. Dotted black line, mean concentration in healthy volunteers; grey shading, 95% CI in healthy volunteers. SD, standard deviation; PBMC, peripheral blood mononuclear cells; LOF, loss-of-function (nonsense, frameshift, or null variant); CI, confidence interval; IL, interleukin; LPS, lipopolysaccharide; ELISA, enzyme-linked immunosorbent assay.

### HA20-causal missense variants are prevalent and associated with immune phenotypes

In AoU, UKBB, and gnomAD, most Pittsburgh cohort *TNFAIP3* missense variants were individually rare (MAF <0.01%), except c.1033T>C (p.Tyr345His, MAF 0.012-0.02%). Collectively, Pittsburgh cohort variants yielded a U.S. prevalence of 1:2,609 (95% CI 1:2,234-3,047; AoU v8) and a global prevalence of 1:3,348 (95% CI 1:2,952-3,799; gnomAD), with the majority attributable to c.1033T>C (p.Tyr345His), leading us to focus on this variant for downstream analysis (**Supp. Table 3)**. Meta□PheWAS of these variants across AoU and UKBB revealed no significant associations, although multiple immune/inflammatory phenotypes trended towards association (aOR 5-15; **Supp. Fig. 3A-C**). In AoU, carriers of p.Tyr345His (n=79) showed 4-5-fold higher odds of high-burden, multi-system immune disease relative to matched controls (n=237) (9+ and 10+ conditions, **Fig. 4A**). p.Tyr345His carriers also had a 31% higher incidence rate of HA20-reported immune/inflammatory diagnoses (IRR: 1.31, 95% CI [1.02, 1.69], p=0.031; **Supp. Fig. 3D**). Accordingly, PSEA enrichment was significant (NES = 1.48, FDR=0.016) suggesting an increased collective burden of inflammatory disease at a population-scale and consistent with the atypical, diffuse phenotypes conferred by hypomorphic missense variants (**Fig. 4B, Supp. Table 2**). Together, these data support the hypothesis that *TNFAIP3* hypomorphic missense variants confer “intermediate” risk of immune/inflammatory disease, lower than full LOF but substantially higher than common risk alleles.

**Figure 4.**
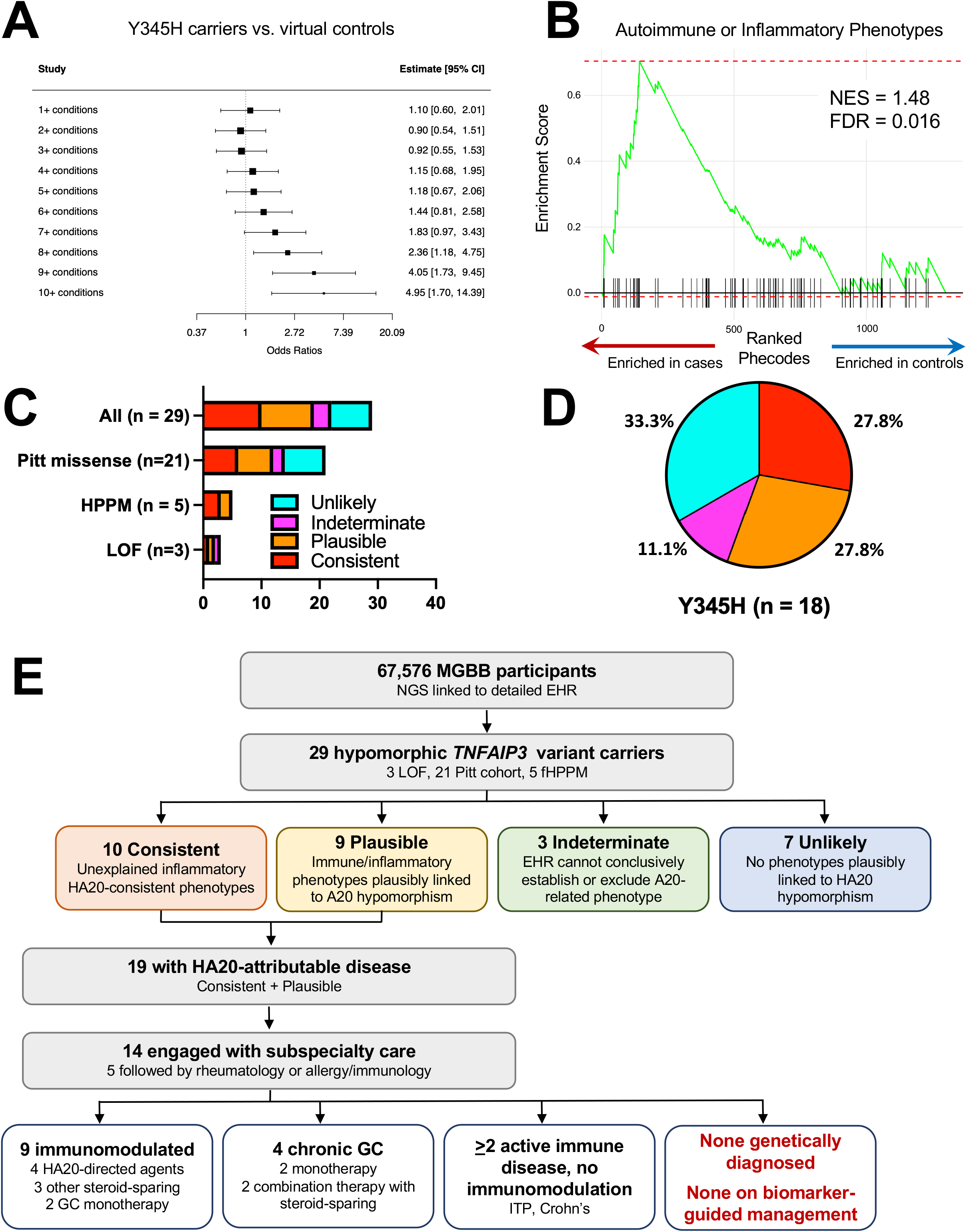
**A.** Forest plot shows odds ratio of having ≥1 HA20-related diagnosis (Fisher’s exact test) at each cumulative threshold in individuals from AoU with linked EHR data and p.Tyr345His, a pathogenic missense *TNFAIP3* variant originally identified in the Pittsburgh cohort (n=79) vs. sex- and age-matched controls (n=237). Black square, each threshold effect size estimate; size of square, weight based on precision; whiskers, 95% CI for each estimate. **B.** PSEA plot shows enrichment of immune/inflammatory phecodes (NES = 1.48, FDR-adjusted p=0.016) in AoU and UKBB carriers of p.Tyr345His (originally identified in the Pittsburgh cohort) relative to other individuals. **C.** Bar graph depicts patients identified in MGB Biobank who harbor pLOF, functionally validated HPPM, or missense variants validated to be pathogenic in the Pittsburgh cohort. In-depth clinical analysis of patient EHRs revealed features that were classified as consistent with (red), plausible (orange), or unlikely (teal) to be HA20-associated conditions. Patients for whom EHR data could not conclusively establish whether a diagnosis was immune-mediated were labeled as indeterminate (pink). **D.** Pie chart depicts percentage of p.Tyr345His carriers (n=18) with features classified in same way as **4C**. **E.** Flowchart of variant identification in the MGB Biobank, patient stratification based on clinical conditions recorded in the EHR, and health services engagement and treatment. Y345H, p.Tyr345His variant in the gene *TNFAIP3*; CI, confidence interval; PSEA, phenotype set enrichment analysis; NES, normalized enrichment score; Phecode, phenotype code; AoU, All of Us; UKBB, United Kingdom Biobank; MGB, Mass General Brigham; pLOF, predicted loss-of-function; HPPM, highly predicted pathogenic missense (with functional validation); EHR, electronic health record.

### Clinical biobank analysis reveals under-recognized, actionable HA20

While population-scale biobank analysis revealed significant associations, and the Pittsburgh cohort validated missense variant effects, neither directly addressed whether genotype-first identification of HA20 and related IEI-HI disorders is clinically actionable. We therefore interrogated the Mass General Brigham (MGB) Biobank, in which 67,576 individuals were prospectively enrolled and underwent next-generation sequencing linked to detailed EHR. As in population biobanks, individuals with pathogenic *TNFAIP3* variants were unexpectedly common: 3 with pLOF variants, 21 with hypomorphic Pittsburgh cohort variants (18 with c.1033T>C, p.Tyr345His), and 14 with HPPM variants (5 fHPPM).

Combining all individuals with hypomorphic variants (n = 29), 10 individuals (34.5%) had otherwise unexplained phenotypes consistent with undiagnosed HA20; 9 additional carriers (31.0%) carried multiple immune/inflammatory diagnoses plausibly related to A20 hypomorphism. For 3 individuals (10.3%), EHR data could neither conclusively establish nor exclude a role for A20 deficiency; 7 individuals (24.1%) had no EHR features consistent with A20-hypomorphic disease (**Fig. 4C-E, Supp. Data 10**). HA20-consistent and HA20-plausible phenotypes were observed across all variant classes: pLOF carriers (1/3 HA20-consistent, 1/3 HA20-plausible), hypomorphic HPPM carriers (3/5 HA20-consistent, 2/5 HA20-plausible), and Pitt missense variant carriers (6/21 HA20-consistent, 6/21 HA20-plausible). Even carriers of c.1033T>C (p.Tyr345His) – a variant so common that it is generally thought too prevalent to cause a rare monogenic disease – had a 55.6% frequency of HA20-consistent or HA20-plausible diagnoses (**Fig. 4D**), consistent with PSEA results. No *TNFAIP3* variant carriers had undergone clinical genetic testing for germline IEI. This even included the pLOF HA20-consistent carrier, who suffered from refractory seropositive rheumatoid arthritis, T-cell large granular lymphocytic leukemia, recurrent non-infectious fevers, pyoderma gangrenosum, and cutaneous lymphocytic vasculitis for years until started on a JAK inhibitor.

Of 19 carriers with consistent or plausible HA20-attributable disease, 14 were engaged with subspecialty care (**Figure 4E, Supp. Data 10**). Of those, 5 were followed longitudinally by rheumatology or allergy/immunology, the specialties most likely to recognize HA20. Nine patients had received immunomodulatory therapies, none informed by a molecular diagnosis or guided by HA20-associated biomarkers (**Fig. 4E**). Instead, patients were treated according to standard algorithms for their individual phenotypically-defined diseases. Four patients had been treated with agents used in HA20-directed algorithms^35^, three had received other steroid-sparing immunomodulators, and two had received glucocorticoid (GC) monotherapy. Four patients required chronic GC, a strategy that remains common in autoimmune diseases despite guideline-based preferences for GC-sparing therapy – at least partly due to the lack of molecular signatures to guide targeted treatment^36^. At least 2 additional carriers had active immune-mediated disease managed without immunomodulation. At least 10 carriers had family members with potentially relevant yet unevaluated diagnoses, including immune-mediated disease, aneurysms, and premature cardiovascular disease.

Together, these results demonstrate that, within a clinical population, most non-preselected *TNFAIP3* hypomorphic variant carriers had phenotypes consistent with or plausibly related to A20, suggesting a penetrance of >50% in this population. Moreover, these findings demonstrate that existing institutional biobanks, many of which have already enrolled tens of thousands of patients at academic medical centers^37^, can identify patients harboring actionable, disease-causal *TNFAIP3* variants. Because established pipelines for returning actionable results already exist within the MGB Biobank^38^, clinical implementation appears feasible at the institutional level, offering a path to targeted treatment for heretofore undiagnosed monogenic disease.

## Discussion

Monogenic diseases are generally assumed to be rare in human populations: even the most common and incompletely penetrant haploinsufficient IEI have been reported in only ∼400-500 individuals globally^39^. However, these assumptions rely on case series rather than systematic epidemiological studies, making it impossible to reliably estimate disease prevalence. While some recent studies have suggested higher-than-expected prevalence of pathogenic variants in population databases, definitive evidence linking variant burden to clinical disease burden has been lacking. We found that IEI-HI genes harbored more predicted deleterious variants in population databases than expected from constraint metrics or published case counts. Most of these genes were strongly and significantly associated with individual immune phenotypes and with collective disease burden at a population level, suggesting that these deleterious variants are not only collectively more common than expected but that they are driving a substantial burden of as-yet unrecognized genetic disease.

To more rigorously test whether this discrepancy reflects true underdiagnosis of clinical disease, we focused on *TNFAIP3*, where loss-of-function variants cause HA20 with reportedly complete penetrance. Despite fewer than 200 cases worldwide, we identified deleterious *TNFAIP3* variants at prevalences as high as 1:24,000, over 100-fold higher than expected. Using unbiased phenome-wide analyses of EHR data, we found that hypomorphic variants significantly increased the odds of immune-mediated and inflammatory conditions. These associations were further supported by deep phenotyping of HA20 patients with missense variants in our referral-based cohort. Functional validation confirmed that identified variants impair A20 function through loss of protein activity, reduced expression, or both. These findings suggest a substantial recognition gap for HA20 and likely for other IEI-HI diseases.

Rare missense variants have previously been reported to cause HA20, but the molecular mechanisms are poorly defined^40–42^. Moreover, some variants reported as disease-driving are classified as benign by ACMG criteria ^43^. Our results help resolve these contradictions by showing that missense *TNFAIP3* mutations are associated with an “intermediate” immunologic phenotype, with inflammatory biomarkers attenuated relative to pLOF variants but elevated compared to healthy individuals. Functional testing suggests that pLOF alleles uniformly impair A20 function and basal expression, while missense variants exhibit heterogeneous mechanisms; some behave like pLOF alleles, others exhibit weakly hypomorphic function, and still others reduce A20 expression despite near-normal NF-κB suppression. Consistent with these molecular findings, deleterious missense variants are associated with a more diffuse and less specific spectrum of immune disease than LOF variants. Although effect sizes are lower for missense variants than for LOF variants, they far exceed the effect sizes of common risk alleles. Moreover, in a hospital-based population, >50% of carriers for the relatively common missense variant c.1033T>C (p.Tyr345His; MAF 0.012-0.02%) have phenotypes plausibly attributed to HA20. The further observation that individuals with deleterious missense variants can develop clinically significant dominantly inherited familial immune dysregulation suggests that such patients should be monitored closely, with a low threshold to consider therapies targeting A20-regulated pathways.

A central challenge in rare variant research has been insufficient power to detect disease associations in population databases. Prior studies have addressed this by analyzing variants with higher allele frequencies (up to 2%) or by restricting analyses to prespecified phenotypes^3^. By collapsing individuals with ultra-rare predicted deleterious variants across multiple alleles, we enabled unbiased phenome-wide testing that revealed large effect sizes for multiple immune diagnoses. Clinician-guided phenotyping in AoU and deep immunophenotyping in our referral cohort provided orthogonal validation. The convergence of evidence across independent data sources demonstrates the power of rare variant collapsing approaches for diseases where individual variants are too rare for traditional association studies. Future studies should explore whether similar patterns exist for diseases caused by other genetic mechanisms.

Our findings have immediate clinical implications. Most importantly, our clinical biobank findings open a pathway to direct actionability through callback amendments to allow return-of-results for confirmatory testing and improved treatment. Such amendments have been successfully implemented across large biobanking consortia to return polygenic risk scores^37^ and within the MGB Biobank for actionable genomic findings^38^. Identification of potentially affected family members suggests that cascade testing could extend this impact. Building on our findings, a return-of-results protocol for IEI is under evaluation at Mass General Brigham, to identify patients with as-yet undiagnosed and undertreated disease. Outside of this immediate actionability, patients presenting with early-onset, multi-system, or treatment-refractory autoimmune and autoinflammatory disease should be considered for genetic testing, especially if they have features of Behcet’s disease or unusual/atypical clinical presentations. For patients with confirmed deleterious variants, proactive monitoring for disease development or progression is warranted. This includes patients with hypomorphic missense variants, who are at high risk of clinically significant disease despite attenuated biomarker profiles. Results within the Pittsburgh referral cohort demonstrate that targeting A20-regulated pathways, including

TNF-NF-κB, inflammasome-IL-1β, and IFN-JAK-STAT, can improve patient outcomes^4,8,35,44^. Because widely available commercial genetic testing panels already incorporate *TNFAIP3*, expanded genetic testing can readily identify individuals who would benefit from precision approaches^4,8,44^.

The implications of these findings extend beyond HA20. Recent work has identified monogenic disease in 1 to 3% of patients diagnosed with common inflammatory conditions like inflammatory bowel disease and multiple sclerosis, challenging the perception that monogenic conditions are ultra-rare and raising critical questions about prevalence and penetrance at a population level ^1,45^. Our study addresses these questions directly, demonstrating higher than expected numbers and disease risk associations for most known haploinsufficiency IEI genes.

This gap between known and estimated disease prevalence may reflect ascertainment bias, poor access to genetic testing, or phenotypic heterogeneity: in HA20, patients may present with isolated autoimmune conditions rather than canonical HA20 phenotypes, leading to misclassification as a common polygenic disease. Our findings suggest that rare IEI-HI variants may act as high-penetrance drivers or modify disease severity within ostensibly polygenic disease populations.

The heterogeneity and frequency of high-burden, multi-system immune disease in *TNFAIP3* variant carriers suggests A20-related diseases may be particularly heterogeneous. This may relate to A20’s diverse regulatory functions, including ubiquitin-editing of the NF-κB, NLRP3, and type I interferon signaling pathways^44,46–49^. Variable expressivity – whether from mutation type (pLOF vs. missense), genetic or environmental modifiers, somatic reversion, alternative promoter usage, or monoallelic exclusion – does not diminish clinical relevance but rather underscores the need for broader genetic testing in patients with unexplained immune dysregulation ^7,50,51^. Identifying the factors modulating expressivity and updating frameworks to account for phenotypic variability should be future priorities.

Our study has several limitations. The referral HA20 cohort is subject to ascertainment bias and may not include individuals with incompletely penetrant variants or mild disease. Conversely, EHR-based phenotyping used for population-wide analyses may be constrained by differences in site-specific data collection and coding practices^52^. Moreover, AoU and UKBB exclusively include adult individuals, potentially limiting our ability to capture patients who may have developed early-onset severe disease. Broad IEI-HI analyses relying on *in silico* predictors may imperfectly capture variant deleteriousness: >50% of HPPM variants were non-hypomorphic, and including benign or weakly hypomorphic variants could dilute observed associations. A20 functional assays, while gold-standard in translational research settings, are not standardized for clinical use^4,8,40^. Variants identified in population databases could not be assessed for baseline A20 expression because patient-derived samples were not available. Hence, it is possible that some excluded, functionally “normal” *TNFAIP3* HPPM variants could have been deleterious, as seen for p.Gly583Glu. Other variants could have been excluded based on computational prediction (e.g. c.2269C>A, p.Pro757Thr) or allele frequency (e.g. c.1033T>C, p.Tyr345His).

We mitigated these issues through stringent HPPM criteria, orthogonal phenotyping, domain-specific analyses, and concordant functional testing, but residual bias remains possible. High throughput screens of *TNFAIP3* function are needed to define the spectrum of A20 hypomorphism and variant pathogenicity at a population level.

We could not exclude deleterious variants in other immune genes in deleterious *TNFAIP3* variant carriers or controls. However, we confirmed the absence of co-occurring pLOF variants in other IEI-haploinsufficiency genes in both cohorts. Other rare, incompletely penetrant variants, if present, would be expected to distribute comparably across groups. Future work integrating functional validation, allele-specific expression studies, gene regulatory networks, and deep clinical phenotyping in large non-referral-based cohorts will be needed.

More broadly, existing analytical frameworks are optimized for extremes of allele frequency and risk effects: common, low-effect size variants for conventional GWAS/PheWAS and ultra-rare, extreme effect size variants for collapsing rare-variant analyses. Moderately rare, moderately penetrant variants like c.1033T>C (p.Tyr345His) fall between these extremes, leading to inconclusive results when existing statistical frameworks are applied. Dedicated approaches to estimate the effects of such intermediate-frequency, moderate-effect variants are needed to reliably identify, monitor, and risk-stratify carriers^53^.

In summary, our findings integrate population genetics, functional validation, and clinical phenotyping to implicate *TNFAIP3* hypomorphic variants as a substantially underrecognized driver of human disease, likely reflecting a broader pattern across haploinsufficiency immune diseases. Expanded genetic testing, updated variant classification frameworks, and comprehensive clinical phenotyping may advance both the mechanistic understanding of monogenic diseases and the ability to diagnose affected individuals. From a clinical standpoint, these approaches provide a concrete pathway to improved outcomes via return-of-results from existing biobanks, broad genetic testing, and pathway-directed therapies in tens of thousands of individuals with hypomorphic variants in *TNFAIP3* and other immune signaling genes.

### AI disclosure

The authors used the University of Pittsburgh Claude for Education AI tool for editorial assistance during manuscript review, including text condensing, consistency and cross-reference checking and formatting review. All scientific content, data analysis, interpretation, methodological choices, and conclusions were performed by the authors, who reviewed and verified all AI-assisted edits and take full responsibility for the accuracy and integrity of the final manuscript.

### Data sharing

Data from gnomAD, UK Biobank, and All of Us are publicly available through their respective portals. The AoU Resource Access Board granted an exemption to the Data and Statistics Dissemination Policy for Table 1. Individual-level clinical data from the University of Pittsburgh referral cohort are not publicly available due to patient privacy; de-identified data may be available from the corresponding author (DMS) upon reasonable request and institutional approval.

### Other Disclosures

Dr. Schwartz has performed consultancy for Sobi and Electra Therapeutics, unrelated to this work. Dr. Sparks has received research support from 10x Genomics, Boehringer Ingelheim, Bristol Myers Squibb, Johnson & Johnson, and Sonoma Biotherapeutics unrelated to this work. He has performed consultancy for AbbVie, AstraZeneca, Boehringer Ingelheim, Bristol Myers Squibb, First Tracks, GSK, Immunovant, Invivyd, Johnson & Johnson, Novartis, Pfizer, Sun, and UCB unrelated to this work. The funders had no role in the decision to publish or preparation of this manuscript. The content is solely the responsibility of the authors and does not necessarily represent the official views of Harvard University, the University of Pittsburgh, affiliated academic health care centers, or the National Institutes of Health.

## Data Availability

Data from gnomADv4, UK Biobank, and All of Us are publicly available through their respective portals. Individual-level clinical data from the University of Pittsburgh referral cohort are not publicly available due to patient privacy; de-identified data may be available from the corresponding author (DMS) upon reasonable request and institutional approval.

https://allofus.nih.gov

https://www.ukbiobank.ac.uk

## Acknowledgements

The authors would like to thank AoU, UKBB, and gnomAD participants. This study used data from the AoU Controlled Tier Dataset v8, available to authorized users on the Researcher Workbench, gnomAD v4, and UK BioBank, available to authorized users on the UKB-Research Analysis Platform. The authors would like to thank Dr. Xiao Peng and Dr. Shmuel Muallem for constructive feedback.

## Funding

This work was supported by NIAID (T32-AI074490, F30-AI200161, R01-AI189495), NIGMS (T32-GM144300), NINDS (R01-NS134651), NIAMS (R01AR080659, R01AR077607, P30 AR070253, 5T32AR007530-42, and P30 AR072577), NHLBI (grant number R01 HL155522), the Jeffrey Modell Foundation, Rheumatology Research Foundation, Samuel and Emma Winters Foundation, University of Pittsburgh Competitive Medicine Research Fund, Sobi, Eli Lilly, the Arthritis Foundation, the R. Bruce and Joan M. Mickey Research Scholar Fund, and the Gordon and Llura Gund Foundation. The Mass General Biobank was supported by Clinical Translational Science Award 1UL1TR002541-01 to Harvard University and Brigham and Women’s Hospital from the National Center for Research Resources.

## Notes

### Competing Interest Statement

Competing interests: All authors have completed the ICMJE uniform disclosure form at www.icmje.org/coi_disclosure.pdf and declare: DMS has received research grants from Eli Lilly and Sobi for the submitted work; DMS has received research grants and honorariums from the Jeffrey Modell Foundation, Rheumatology Research Foundation, Samuel and Emma Winters Foundation, and the Arthritis Foundation; JAS has received grants from the R. Bruce and Joan M. Mickey Research Scholar Fund, and the Gordon and Llura Gund Foundation; no other relationships or activities that could appear to have influenced the submitted work.

### Author Declarations

Individuals in the paper were referred for evaluation under University of Pittsburgh Institutional Review Board approved STUDY22060106; all participants provided written informed consent.

### Summary of Updates

We have conducted additional studies to extend the generalizability of the paper, including data on other haploinsufficiency diseases and from the Mass General Biobank. We have updated our funding sources based on new collaborations and funding.

