## Supplementary figures and images for "How Rare is Rare? *TNFAIP3* Variants and the High Collective Burden of Haploinsufficiency"

### Supplementary Figures

#### Supplementary Figure 1

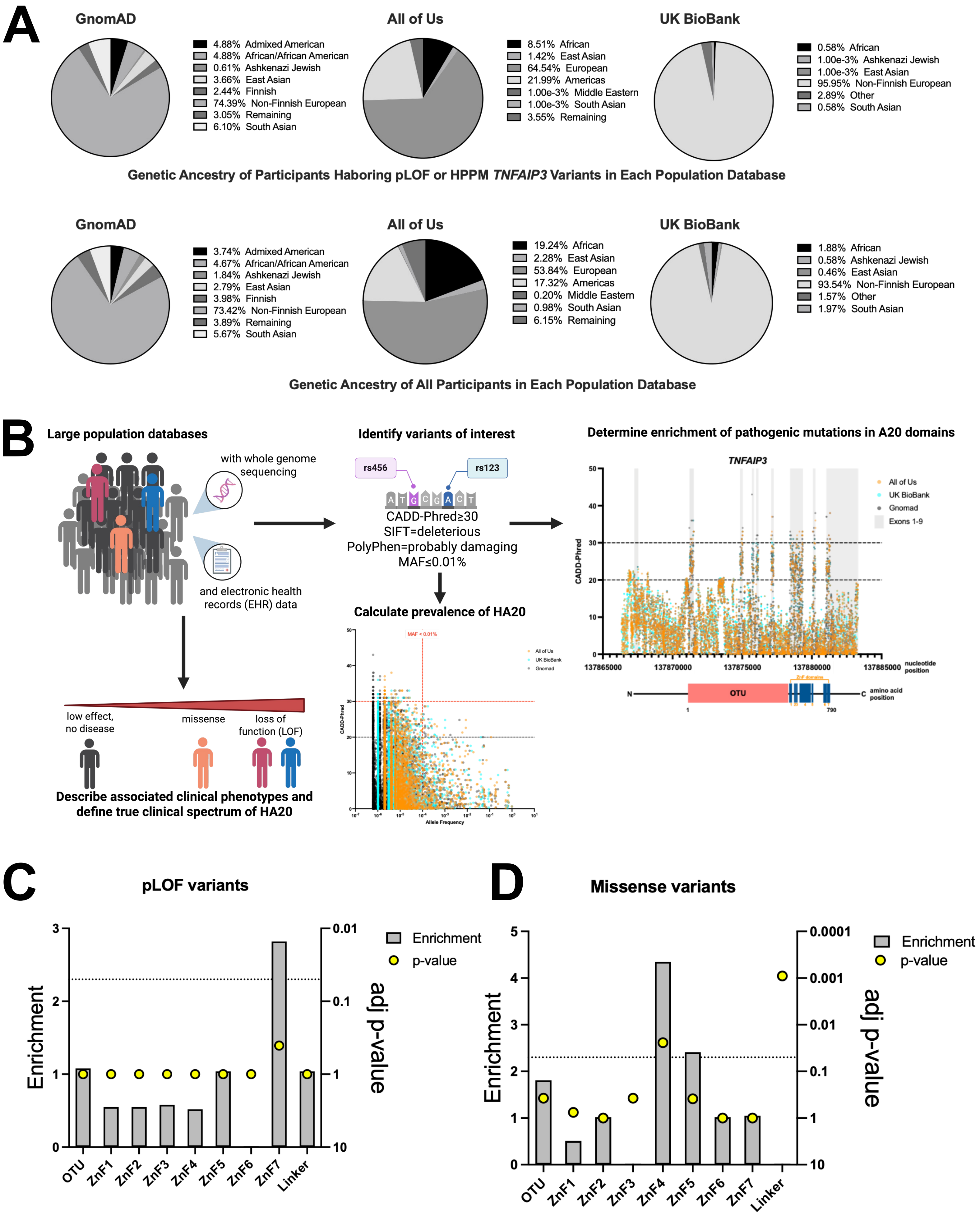

#### Supplementary Figure 2

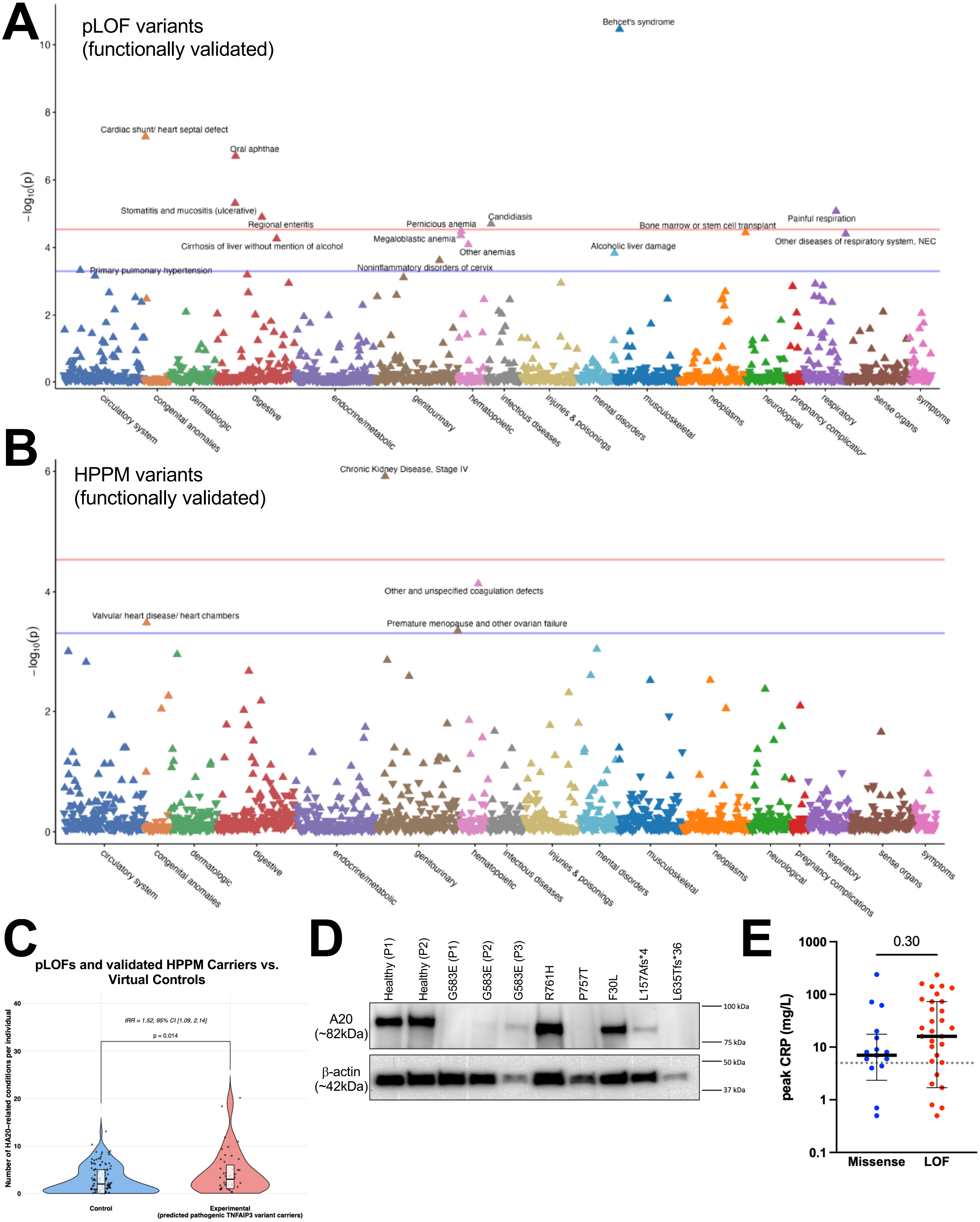

#### Supplementary Figure 3

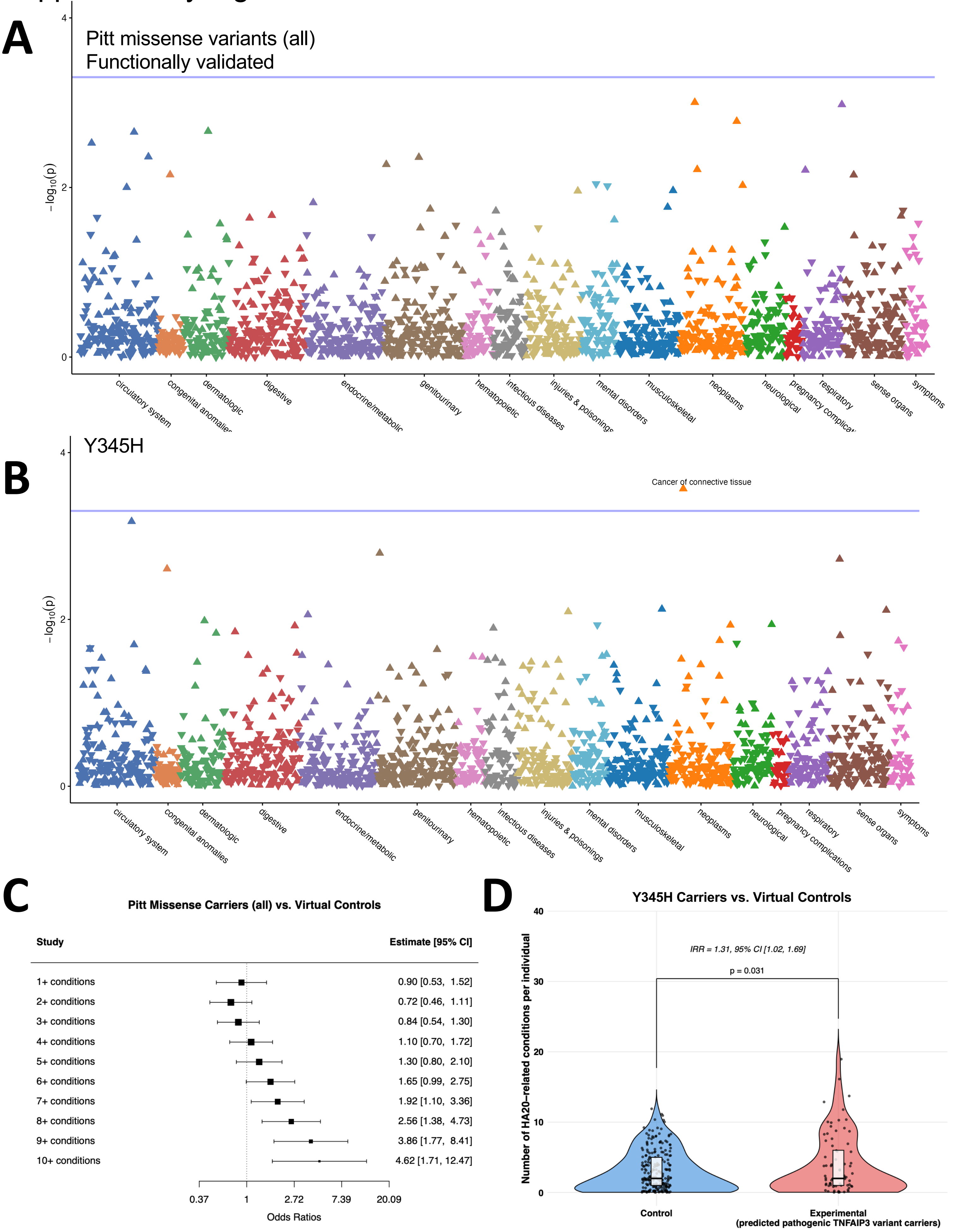
