## Supplementary Figure Legends for "How Rare is Rare? *TNFAIP3* Variants and the High Collective Burden of Haploinsufficiency"

### Supplementary Figure Legend

**Supplementary Figure S1. A.** Genetic ancestry of all participants harboring pLOF or HPPM variants in population databases compared to genetic ancestry of all participants in each population database. There was no significant enrichment for certain ancestries in individuals harboring pathogenic *TNFAIP3* variants. Genetic ancestry for all individuals in each population database is inferred using principal component analysis and is publicly available data. Percentages (%) are shown next to each ancestry for each pie chart. **B.** Schematic shows identification pipeline for pLOF or HPPM *TNFAIP3* variants in three large population databases based on minor allele frequency (MAF) and pathogenicity prediction tools (CADD-Phred, SIFT, and PolyPhen). Variants of interest were plotted by amino acid position to calculate enrichment of pLOF vs. HPPM variants in each A20 functional domain. Individuals with variants of interest who also had associated electronic health records data were included in subsequent meta-PheWAS analysis or manual comparative clinical phenotyping. **C,D.** Bar graphs show enrichment scores and p-values (Fisher's exact test, Benjamini-Hochberg correction) for unique pLOF (C) and HPPM (D) population variants in each A20 domain. Bars, enrichment score; Yellow circle, p-value; Dotted line, adjusted p=0.05 (significance threshold); pLOF, predicted loss of function; HPPM high predicted pathogenic missense; CADD-Phred, Combined Annotation Dependent Depletion, Phred-Scaled; SIFT, Sorting Intolerant From Tolerant; PolyPhen, Polymorphism Phenotyping; PheWAS, phenome wide association study; AoU, All of Us; UKBB, UK Biobank; gnomAD, Genome Aggregation Database.

**Supplementary Figure S2. A,B.** Manhattan plot shows meta-PheWAS (AoU and UKBB) for individuals with (A) pLOF *TNFAIP3* variants only or (B) functionally validated HPPM (fHPPM) variants only. Each data point represents a phecode; color delineates its phecode category. Red line, threshold for statistical significance based on Bonferroni adjustment ($p < 2.75\times10^{-5}$); blue line, suggestive threshold ($p < 5\times10^{-4}$). **C.** Comparison of total number of HA20-related conditions per individual and incidence rate ratio between experimental cases (red violin plot, n=43) and virtual controls (blue violin plot, n=129) using negative binomial regression (two-sided). Within each violin, individual data points are overlaid. The white box denotes the interquartile range, with the median indicated by the black horizontal line. Cases are individuals in AoU with pLOF and functionally validated HPPM *TNFAIP3* variants and linked EHR data; controls are sex- and age-matched and harbor only common, benign *TNFAIP3* variants. **D.** Representative western blot showing basal expression of A20 and β-actin control from healthy control volunteers and patient PBMCs. Patients include two individuals with HA20 due to pLOF variants (Leu157Alafs*4, Lys635Thrfs*36), four individuals with pathogenic missense variants by ACMG criteria (Gly583Glu [n = 3]; Pro757Thr [n = 1]), and two individuals with missense variants characterized as VUS by ACMG criteria (Arg761His, Phe30Leu). **E.** peak CRP, mg/L. Dashed line denotes reference range cutoff for clinical assay (0-5 mg/L). pLOF, predicted loss of function; HPPM high pathogenic predicted missense; PheWAS, phenome wide association study; Phecode, phenotype code; AoU, All of Us; UKBB, UK Biobank; GnomAD, Genome Aggregation Database; PBMCs, peripheral blood mononuclear cells; VUS, variant of uncertain significance.

**Supplementary Figure S3. A.** Manhattan plot shows meta-PheWAS (AoU and UKBB) for carriers of functionally validated missense variant identified in the Pittsburgh cohort. **B.**

Manhattan plot shows meta-PheWAS (AoU and UKBB) for carriers of the pathogenic p.Tyr345His variant, identified in the Pittsburgh cohort. Each data point represents a phecode; color delineates its phecode category. Red line, Bonferroni-corrected threshold for statistical significance ($p<2.75\times10^{-5}$); blue line, suggestive threshold ($p <5\times10^{-4}$). **C.** Forest plot shows odds ratio of having ≥1 HA20-related diagnosis (Fisher's exact test) at each cumulative threshold in individuals from AoU with linked EHR data and missense variants validated to be pathogenic from the Pittsburgh cohort (n=107) vs. sex- and age-matched controls (n=321). Black square, each threshold effect size estimate; size of square, weight based on precision; whiskers, 95% CI for each estimate. PheWAS, phenome wide association study; AoU, All of Us; UKBB, UK Biobank; Phecode, phenotype code; CI, confidence interval. **D.** Violin plots show comparison of total number of HA20-related conditions per individual and incidence rate ratio between experimental cases (red, n=79) and virtual controls (blue, n=237) using negative binomial regression (two-sided). Within each violin, individual data points are overlaid. The white box denotes the interquartile range, with the median indicated by the black horizontal line. Cases are individuals in AoU who harbor the p.Tyr345His variant and have linked EHR data; controls are sex- and age-matched and only harbor common, benign *TNFAIP3* variants.
