## Supplementary Table 1 for "How Rare is Rare? *TNFAIP3* Variants and the High Collective Burden of Haploinsufficiency"

**Supplementary Table 1. Summary data, HA20 patients with missense variants, University of Pittsburgh cohort**

| **Patient** | **Family** | **Sex** | **Age at onset** | **Age at diagnosis** | **Variant** | **Inheritance (if known)** | **MAF (gnomad v4)** | **Predictor scores** | **NF-κB suppression** | **Basal A20 expression** | **ACMG classification and why** | **Main clinical features** | **Acute phase reactants, immunologic features, lab abnormalities** | **Initial treatment** | **Most effective treatment** |
| --- | --- | --- | --- | --- | --- | --- | --- | --- | --- | --- | --- | --- | --- | --- | --- |
| P1 | 1 | M | 0-5 years | 5-10 years | NM_001270508.2 c.1033T>C, p.Tyr345His | Paternal | Overall: 0.0001246, PopMAX: 0.0001761 | CADD Phred 28.6, Alphamissense 0.985, Revel 0.38, SIFT deleterious, PolyPhen probably damaging | Strong LOF | Normal expression | LP: PP1 (segregation) + PS3 (functional) + PM2 (population) | Recurrent upper respiratory infection, severe asthma, chronic recurrent multifocal osteomyelitis, abdominal pain, fevers | Low IgM, eosinophilia, (+) ANA, elevated CRP | CS, TNF inhibitors | IL1R antagonist |
| P2 | 1 | M | 0-5 years | 10-15 years | NM_001270508.2 c.1033T>C, p.Tyr345His | Paternal | Overall: 0.0001246, PopMAX: 0.0001761 | CADD Phred 28.6, Alphamissense 0.985, Revel 0.38, SIFT deleterious, PolyPhen probably damaging | Strong LOF | Normal expression | LP: PP1 (segregation) + PS3 (functional) + PM2 (population) | Oral ulcers, fevers, arthralgia, abdominal pain, cystic acne | Low IgM | CS, Colchicine | IL1R antagonist |
| P3 | 1 | M | early childhood | 35-40 years | NM_001270508.2 c.1033T>C, p.Tyr345His | Suspect maternal | Overall: 0.0001246, PopMAX: 0.0001761 | CADD Phred 28.6, Alphamissense 0.985, Revel 0.38, SIFT deleterious, PolyPhen probably damaging | Strong LOF | Normal expression | LP: PP1 (segregation) + PS3 (functional) + PM2 (population) | Oral ulcers, fevers, arthralgia, abdominal pain | Low C4 | CS, Colchicine, PDE4 inhibitor | IL1R antagonist |
| P4 | 2 | F | early childhood | 5-10 years | NM_001270508.2 2 c.1033T>C, p.Tyr345His | Maternal | Overall: 0.0001246, PopMAX: 0.0001761 | CADD Phred 28.6, Alphamissense 0.985, Revel 0.38, SIFT deleterious, PolyPhen probably damaging | Strong LOF | Normal expression | LP: PP1 (segregation) + PS3 (functional) + PM2 (population) | Oral ulcers, Fevers, arthralgia, atopy | IFN signature (+) | Colchicine | Colchicine |
| P5 | 2 | F | 5-10 years | 30-35 years | NM_001270508.2 c.1033T>C, p.Tyr345His | Unknown (incomplete history) | Overall: 0.0001246, PopMAX: 0.0001761 | CADD Phred 28.6, Alphamissense 0.985, Revel 0.38, SIFT deleterious, PolyPhen probably damaging | Strong LOF | Normal expression | LP: PP1 (segregation) + PS3 (functional) + PM2 (population) | Oral and genital ulcers | IFN signature (+) | None | Colchicine |
| P6 | 3 | F | 0-5 years | 15-20 years | NM_001270508.2 c.1748G>A, p.Gly583Glu | Maternal | Not in gnomad | CADD Phred 20.6; Alphamissense 0.106, Revel 0.11, SIFT tolerated, PolyPhen benign | Normal | Low expression | LP: PP5 (reputable source) + PS3 (functional/expression) + PM2 (population) | Recurrent upper respiratory infection, oral ulcers, conjunctival erythema, myalgia, arthralgia, lymphadenopathy, abdominal pain with colonic lymphoid hyperplasia | ANA (+), lymphopenia, low IgM, poor vaccine responses, high fecal calprotectin | Colchicine,-IL-1β antibody | IL1R antagonist + colchicine |
| P7 | 3 | F | early childhood | 45-50 years | NM_001270508.2 c.1748G>A, p.Gly583Glu | Paternal | Not in gnomad | CADD Phred 20.6; Alphamissense 0.106, Revel 0.11, SIFT tolerated, PolyPhen benign | Normal | Low | LP: PP5 (reputable source) + PS3 (functional/expression) + PM2 (population) | Oral ulcers, arthralgia, abdominal pain, sicca, salivary gland swelling | ANA (+), lymphopenia, low IgA, poor vaccine responses | Colchicine | IL1R antagonist + colchicine + hydroxy-chloroquine |
| P8 | 3 | M | early childhood | 85-90 years | NM_001270508.2 c.1748G>A, p.Gly583Glu | Unknown | Not in gnomad | CADD Phred 20.6; Alphamissense 0.106, Revel 0.11, SIFT tolerated, PolyPhen benign | Normal | Low | LP: PP5 (reputable source) + PS3 (functional/expression) + PM2 (population) | Oral ulcers, abdominal pain, pleural effusions | ANA (+), RF (+), SSB(+), CD4 lymphopenia, elevated CRP | Colchicine | IL1R antagonist + colchicine |
| P9 | 4 | M | 0-5 years | 0-5 years | NM_001270508.2 c.1748G>A, p.Gly583Glu | Maternal | Not in gnomad | CADD Phred 20.6; Alphamissense 0.106, Revel 0.11, SIFT tolerated, PolyPhen benign | Normal | Low | LP: PP5 (reputable source) + PS3 (functional/expression) + PM2 (population) | Recurrent acute otitis media infections responsive to antibiotics; noninfectious recurrent fever, tonsillitis, lymphadenopathy, rash, abdominal pain, orogenital ulcers | Low C4, high CRP, high sIL2R, CD8+ lymphocytosis, poor vaccine responses | Colchicine | IL1R antagonist |
| P10 | 4 | F | early childhood | 30-35 years | NM_001270508.2 c.1748G>A, p.Gly583Glu | Unknown (suspect maternal) | Not in gnomad | CADD Phred 20.6; Alphamissense 0.106, Revel 0.11, SIFT tolerated, PolyPhen benign | Normal | Low | LP: PP5 (reputable source) + PS3 (functional/expression) + PM2 (population) | Recurrent fever, tonsillitis, oral ulcers, rash, sicca, inflammatory arthritis, diarrhea, Raynaud's, ITP | Elevated transaminases, low IgM, (+) S100A8/9 | Colchicine | Colchicine |
| P11 | 5 | F | 0-5 years | 30-35 years | NM_001270508.2 c.2269C>A, p.Pro757Thr | Maternal | Overall: 0.000009927, PopMAX: 0.00001603 | CADD Phred 9.8, Alphamissense 0.06, Revel 0.03, SIFT tolerated, PolyPhen benign | Strong LOF | Low | LP: PS3 (functional) + PM2 (population) | Recurrent upper respiratory infections, pleuritis, oral ulcers, bladder ulcers, inflammatory bowel disease, duodenal mucosal atrophy, sicca, small fiber neuropathy, psoriasiform rash, axial spondyloarthritis | Poor vaccine responses, high CRP, high S100A8/9, high S100A12, high fecal calprotectin | CS, Colchicine, Azathioprine, TNF inhibitors, PDE4 inhibitors, Sulfasalazine | TNF inhibitor + Sulfasalazine + Methotrexate |
| P12 | 6 | M | 0-5 years | 0-5 years | NM_001270508.2 c.2269C>A, p.Pro757Thr | Maternal | Overall: 0.000009927, PopMAX: 0.00001603 | CADD Phred 9.8, Alphamissense 0.06, Revel 0.03, SIFT tolerated, PolyPhen benign | Strong LOF | Low | LP: PS3 (functional) + PM2 (population) | Recurrent fever, oral ulcers, failure to thrive, lymphadenopathy, eczematous dermatitis, pustular rash | Leukocytosis, CD4+ lymphocytosis, low IgG, poor vaccine responses | CS | Colchicine |
| P13 | 7 | M | 0-5 years | 0-5 years | NM_001270508.2 c. 688G>A, p.Gly230Ser | Paternal | Overall: 0.000001239, PopMAX: 0.000001695 | CADD Phred 26.8, Alphamissense 0.132, Revel 0.25, SIFT tolerated, PolyPhen possibly damaging | Weak LOF | Normal | LP: PP1 (segregation)+ PM3 (functional) + PM2 (population) | Neutrophilic urticarial dermatoses, recurrent fevers, elevated transaminases | ANA (+), high CXCL9, high S100A8/9, high S100A12, lymphocytosis | CS, IL-1b antibody | IL-1β antibody |
| P14 | 7 | M | early childhood | 40-45 years | NM_001270508.2 c. 688G>A, p.Gly230Ser | Unknown (suspect paternal) | Overall: 0.000001239, PopMAX: 0.000001695 | CADD Phred 26.8, Alphamissense 0.132, Revel 0.25, SIFT tolerated, PolyPhen possibly damaging | Weak LOF | Normal | LP: PP1 (segregation)+ PM3 (functional) + PM2 (population) | Childhood-onset type 1 diabetes and autoimmune thyroiditis, severe hyperinflammatory reactions to bee stings | None | None | None |
| P15 | 8 | F | 0-5 years | 15-20 years | NM_001270508.2 c.1504C >T, p.Arg502Trp | Paternal | Overall: 0.00001363, PopMAX: 0.00003124 | CADD Phred 26.2, Alphamissense 0.153, Revel 0.36, SIFT deleterious, PolyPhen possibly damaging | Weak LOF | Normal | LP: PP1 (segregation)+ PM3 (functional) + PM2 (population) | Post-viral hyperinflammatory episodes, aseptic meningitis, oral ulcers, granulomatous gingivitis, aortitis, uveitis, erythema nodosum | high CRP, high CXCL9, high S100A8/9, high S100A12, (+) IFN signature, high IgG, high IgM, | CS, TNF inhibitors | TNF inhibitor + Methotrexate |
| P16 | 8 | M | early childhood | 40-45 years | NM_001270508.2 c.1504C >T, p.Arg502Trp | Unknown (suspect paternal) | Overall: 0.00001363, PopMAX: 0.00003124 | CADD Phred 26.2, Alphamissense 0.153, Revel 0.36, SIFT deleterious, PolyPhen possibly damaging | Weak LOF | Normal | LP: PP1 (segregation)+ PM3 (functional) + PM2 (population) | Recurrent oral ulcers, periodic fevers, cervical lymphadenopathy, psoriasiform rash, inflammatory arthritis, bloody diarrhea | high CRP, high CXCL9, high S100A8/9 | Colchicine | Colchicine |
| P17 | 9 | M | early childhood | 30-35 years | NM_001270508.2 c.877G>A, p.Asp293Asn | Maternal | Not in gnomad | CADD Phred 24.4, Alphamissense 0.097, Revel 0.11, SIFT tolerated, PolyPhen possibly damaging | Weak LOF | Normal | LP: PP1 (segregation)+ PM3 (functional) + PM2 (population) | Recurrent oral ulcers, fever, myalgia, acute inflammatory demyelinating polyneuropathy, lymphadenopathy, autoimmune thyroiditis | Low IgG, SSA positive, elevated CRP, CD4 lymphopenia | CS, IVIG | IL1R antagonist + IL-1β antibody + IVIG |
| P18 | 9 | F | Before age 10 | 60-65 years | NM_001270508.2 c.877G>A, p.Asp293Asn | Unknown (suspect maternal) | Not in gnomad | CADD Phred 24.4, Alphamissense 0.097, Revel 0.11, SIFT tolerated, PolyPhen possibly damaging | Weak LOF | Normal | LP: PP1 (segregation)+ PM3 (functional) + PM2 (population) | Recurrent fevers, oral ulcers, ocular dryness, joint stiffness | None | Colchicine | Colchicine |

PopMAX, maximum frequency within specific subpopulations in gnomad v4; CADD Phred, Phred-scaled Combined Annotation Dependent Depletion; SIFT, Sorting Intolerant From Tolerant; LP, likely pathogenic; CRP, C reactive protein; S100, S100 calcium binding protein; CXCL, CXC-motif ligand; IFN, interferon; Ig, immunoglobulin; CS, corticosteroids; TNF, tumor necrosis factor; IL, interleukin; IVIG, intravenous immunoglobulin
