## Supplementary Table 2-3 for "How Rare is Rare? *TNFAIP3* Variants and the High Collective Burden of Haploinsufficiency"

**And Supplementary Table 2. Clinical features, University of Pittsburgh HA20 cohort**

|  | Missense | LOF | Odds Ratio | 95% CI | p-val (raw) | p-val (adj) |
| --- | --- | --- | --- | --- | --- | --- |
| **DEMOGRAPHICS** | | | | | | |
| Number of patients | 18 | 48 | - |  | - | - |
| Number of families | 9 | 19 | - |  | - | - |
| Mean age of onset | 4 | 6 | - |  | - | - |
| Mean age of diagnosis | 29 | 28 | - |  | - | - |
| **AUTOINFLAMMATION** | | | | | | |
| Oral ulcers | 88.9% | 91.7% | 1.37 | 0.11-10.65 | 0.66 | 1 |
| Genital ulcers | 11.1% | 62.5% | 12.82 | 2.56-127.99 | 2.2E-04 | 3.2E-03 |
| Rash | 83.3% | 68.8% | 0.45 | 0.07-1.93 | 0.35 | 0.86 |
| Arthritis/arthralgia | 77.8% | 77.1% | 0.96 | 0.19-3.99 | 1 | 1 |
| Ocular disease | 38.9% | 43.8% | 1.22 | 0.36-4.4 | 0.79 | 1 |
| IBD-like | 27.8% | 25.0% | 0.87 | 0.23-3.77 | 1 | 1 |
| Vascular disease | 22.2% | 25.0% | 1.16 | 0.29-5.8 | 1 | 1 |
| Recurrent fever | 72.2% | 64.6% | 0.71 | 0.17-2.58 | 0.77 | 1 |
| Serositis | 27.8% | 18.8% | 0.61 | 0.15-2.73 | 0.50 | 0.97 |
| **AUTOIMMUNITY** | | | | | | |
| Autoantibodies | 27.8% | 85.4% | 14.34 | 3.53-69.85 | 1.6E-05 | 4.8E-04 |
| Autoimmune thyroid disease | 5.6% | 16.7% | 3.35 | 0.39-159.49 | 0.43 | 0.88 |
| Type 1 diabetes | 5.6% | 6.3% | 1.13 | 0.08-62.93 | 1 | 1 |
| Autoimmune hepatitis | 0.0% | 16.7% | N/A | N/A | 0.10 | 0.40 |
| Immune kidney disease | 5.6% | 8.3% | 1.54 | 0.14-80.49 | 1 | 1 |
| Lung involvement | 55.6% | 25.0% | 0.27 | 0.07-0.96 | 0.04 | 0.28 |
| Autoimmune cytopenias | 16.7% | 43.8% | 3.82 | 0.91-23.25 | 0.05 | 0.28 |
| CNS inflammatory disease | 38.9% | 14.6% | 0.27 | 0.07-1.13 | 0.04 | 0.28 |
| Peripheral neuropathy | 11.1% | 6.3% | 0.54 | 0.06-7 | 0.61 | 0.98 |
| Sicca symptoms | 22.2% | 10.4% | 0.41 | 0.08-2.38 | 0.24 | 0.78 |
| **LYMPHOPROLIFERATION** | | | | | | |
| Lymphoma | 0.0% | 4.2% | N/A | N/A | 1 | 1 |
| Lymphadenopathy | 33.3% | 41.7% | 1.42 | 0.41-5.44 | 0.58 | 0.98 |
| Splenomegaly | 11.1% | 29.2% | 3.24 | 0.62-32.81 | 0.20 | 0.72 |
| **IMMUNODEFICIENCY** | | | | | | |
| CVID | 11.1% | 4.2% | 0.35 | 0.02-5.26 | 0.30 | 0.79 |
| Low IgG | 11.1% | 12.5% | 1.14 | 0.18-12.7 | 1 | 1 |
| Low IgA | 11.1% | 12.5% | 1.14 | 0.18-12.7 | 1 | 1 |
| Low IgM | 22.2% | 6.3% | 0.24 | 0.03-1.6 | 0.08 | 0.39 |
| **NON-IMMUNE** | | | | | | |
| Developmental delay | 11.1% | 4.2% | 0.35 | 0.02-5.26 | 0.30 | 0.79 |
| Osteoporosis | 0.0% | 8.3% | N/A | N/A | 0.57 | 0.98 |
| Short stature | 5.6% | 16.7% | 3.35 | 0.39-159.49 | 0.43 | 0.88 |

p-val (raw), raw p-values, Fisher’s exact test, p-val (adj), Benjamini-Hochberg adjusted p-values; LOF, loss of function (truncating/nonsense, null including large deletions and microdeletions, frameshift); IBD, inflammatory bowel disease; CNS, central nervous system; CVID, common variable immunodeficiency; Ig, immunoglobulin

**Supplementary Table 3. Prevalence estimates of missense variants (University of Pittsburgh HA20 referral cohort) in the US (AoU), UK (UKBB), and globally (gnomAD)**

Six HA20-causal missense variants were identified within the University of Pittsburgh HA20 cohort. These included p.Gly230Ser, p.Asp293Asn, p.Tyr345His, p.Arg502Trp, p.Gly583Glu, and p.Pro757Thr. Individuals harboring these variants were identified in AoU, UKBB, and GnomAD. Mean prevalence of the variants and affected number of individuals were estimated based on a 2026 U.S. population estimate of 3.49E8, U.K. population estimate of 6.99E7, and a global population estimate of 8.3E9. AoU, All of Us; UKBB, UK Biobank; GnomAD, genome aggregation database.

|  | | GnomAD v4.0 | All of Us v8 | UK BioBank* |
| --- | --- | --- | --- | --- |
| Total # of genomes | | 807,162 | 414,830 | 425,966 |
| # of Pitt HPPM (# subjects) | | 4 (241) | 5 (159) | 3 (194) |
| Mean prevalence of Pitt HPPM (95% CI) | | 1:3,348  (1:2,952-3,799) | 1:2,609  (1:2,234-3,047) | 1: 2,196  (1:1,908-2,527) |
|  | Estimated # affected in U.S. | 104,241 | 133,768 | ----- |
|  | Estimated # affected in U.K. | 20,878 | ----- | 31,831 |
|  | Estimated # affected globally | 2,479,092 | 3,181,296 | 3,779,599 |
| # of p.Tyr345His carriers | | 201 | 116 | 171 |
| Mean prevalence of Tyr345His (95% CI) | | 1:4,015  (1:3,497-4,609) | 1:3,576  (1:2,982-4,289) | 1:2,491  (1:2,145-2,893) |
|  | Estimated # affected in U.S. | 86,924 | 97,595 | ----- |
|  | Estimated # affected in U.K. | 17,410 | ----- | 28,061 |
|  | Estimated # affected globally | 2,067,248 | 2,321,029 | 3,331,995 |

* Due to the UKBB shutdown, prevalence estimates used data only from individuals with whole genome sequencing data *and* electronic health records (EHR) data, whose EHR data mapped to at least one phecode.
