## Supplementary Appendix (Methods) for "How Rare is Rare? *TNFAIP3* Variants and the High Collective Burden of Haploinsufficiency"

**Supplementary Appendix: Patient Histories**

Detailed patient histories are available from the corresponding author (DMS) upon request; medRxiv restrictions preclude their inclusion in this preprint.

**Supplementary Methods**

*Participants and Data Sources*

Whole genome sequencing (WGS) and electronic health record (EHR)–linked data were extracted from the All of Us Research Program (AoU; Controlled Tier v8, n=414,830; January 2018–October 2023) and the UK Biobank (UKBB; n=502,134 as of November, 6th 2024; n=425,966 with linked EHR data used for variant extraction and prevalence calculations; recruitment 2006–2010), together with aggregated population reference data from gnomAD v4.0 (n=807,162; 730,947 exomes and 76,215 genomes, enriched for European ancestry owing to UKBB inclusion)^10-12^. All datasets were aligned to GRCh38. AoU participants (≥18 years, recruited across the United States) and UKBB participants (ages 40–69, recruited in the United Kingdom) were not selected for specific diseases and were linked to EHR data^10^^,^^11^. In contrast, gnomAD aggregates data from multiple cohorts, attempts to exclude individuals with severe pediatric Mendelian disease, and is not EHR-linked^13^. Diagnostic codes (ICD-9/10) were mapped to phecodes using Phecode v1.2 for phenome-wide association studies (PheWAS) and manual phenotyping. Genetic ancestry for individuals with selected *TNFAIP3* variants in AoU, UKBB, and gnomAD was inferred by using principal component analysis to compare genetic similarity of participants to global reference populations (**Supp. Fig. 1A**)^14-17^.

*University of Pittsburgh HA20 Cohort*
Individuals with suspected pathogenic *TNFAIP3* variants, identified by multigene panel and next‑generation clinical sequencing, were referred for evaluation. All participants provided informed consent under University of Pittsburgh IRB‑approved STUDY22060106. Variant classification followed ACMG criteria (allele frequency, predicted deleteriousness, functional testing, and genotype–phenotype cosegregation)^18^. Immunologic profiling included a standardized panel of assays: S100A8/9, S100A12, and IL-18 (ELISA). Type I interferon gene score (NanoString), LPS-induced IL-1β secretion, and basal A20 expression (Western blot) was also assessed^19-22^. Mutant A20 constructs were tested by luciferase assay in A20‑deficient HEK293 cell lines.

*MGB BioBank Cohort*

The MGBB is a prospective institutional electronic health record linked biorepository that includes data from 67,576 participants who have underwent whole exome/genome sequencing. Genomic data from MGBB participants was aligned to hg38 and joint variant calling was performed for the entire cohort using Genome Analysis Toolkit (GATK). Resulting vcf files were then processed using *bcftools.* High-quality variants (FILTER PASS and GQ>20) in the region of *TNFAIP3* were then annotated using variant effect predictor (VEP). Variants in *TNFAIP3* were then classified as described below. EHR of participants carrying pLOF, HPPM, or Pitt cohort missense variants were then reviewed by CAR to determine if participants carried phenotypes consistent with HA20 spectrum disease or plausibly related to A20 hypomorphism. Carriers were classified into four categories based on chart review: **Consistent** (unexplained inflammatory or HA20-consistent phenotypes with no alternative explanation); **Plausible** (multiple immune or inflammatory diagnoses plausibly related to A20 hypomorphism); **Indeterminate** (EHR data could neither conclusively establish nor exclude an A20-related phenotype); and Unlikely (no EHR features plausibly linked to HA20 hypomorphism). De-identified clinical data was additionally reviewed by author DS before a final classification made for each patient. This work was approved and conducted under MGB IRB protocol #2019P000264.

*TNFAIP3 Variant Ascertainment and Annotation*
Genomic data from AoU, UKBB, and gnomAD were quality‑filtered^14^^,^^17^. *TNFAIP3* variants of interest met minor allele frequency (MAF) <0.01% and predicted deleteriousness thresholds (SIFT, PolyPhen, CADD‑Phred v1.6)^23-25^: predicted loss-of-function (pLOF; frameshift, nonsense, or splice-null variants) or high predicted pathogenic missense (HPPM; missense variants with CADD-Phred>30; SIFT=deleterious; PolyPhen=probably damaging) (**Supp. Fig. 1B**). For variants of interest, we analyzed nucleotide position, amino‑acid change, and functional effects. Amino‑acid changes were localized to A20 domains using sequence alignment. HPPM and pLOF variants were quantified per domain and normalized to unique mutations across resources to compute enrichment scores.

$$\frac{\frac{Mutation\left( within domain \right)}{All unique mutations}}{\frac{Amino acids\left( within domain \right)}{All amino acids}}$$

*HA20 Prevalence Analysis*

We calculated the prevalence of pLOF variants (frameshift, nonsense, splice‑null predicted to abrogate function) and HPPM variants (CADD‑Phred ≥30, SIFT=deleterious, PolyPhen=probably damaging, MAF <0.01%, no homozygotes) in each database. U.S. and global counts were extrapolated from 2026 population estimates (3.49E8 for U.S., 6.99E7 for U.K., and 8.3E9 for global). AoU statistics were used for U.S. prevalence estimates, UKBB statistics were used for U.K. prevalence estimates, and GnomAD statistics were used for global prevalence estimates. Code for the variant extractions is provided in the All of Us Researcher Workbench Controlled Tier and UK BioBank Research Analysis Platform (UKB-RAP).

*Comparative Clinical Phenotype Analysis*

Individuals carrying variants of interest (pLOF, HPPM) from the population databases and missense variants from the Pittsburgh HA20 cohort were evaluated. EHR data from AoU supported descriptive analyses, with sex‑ and age‑matched virtual controls (‘MatchIt’ package in R; exact matching on sex, nearest-neighbor matching on age with no caliper, ratio of 3 controls:1 case). Virtual controls (initial pool n=255,035) were required to have EHR data, carry only common (benign, synonymous) *TNFAIP3* variants (MAF >20%), and lack rare (potentially pathogenic) *TNFAIP3* variants (MAF <0.01%). Case/control classification was based solely on *TNFAIP3* genotype; we did not screen for or exclude cases or controls harboring rare variants in other IEI genes. Cases and controls were required to have at least one recorded ICD-9 or ICD-10 condition in the EHR to be included in downstream phenotype analysis. We focused a priori on potentially HA20‑related phenotypes (e.g., oral/genital ulcers, recurrent fever, rash, arthritis, IBD‑like disease, vascular disease, autoantibodies, autoimmune endocrine disease, liver disease, cytopenias, lymphadenopathy, recurrent infections). A permissive threshold was used to capture the full burden of potential HA20-related features (**Supplementary Data 8**).

Number of potentially HA20-related conditions were counted per individual through manual clinician-led AoU EHR review. For each cumulative condition-count threshold (≥1 through ≥10 HA20-related conditions), the odds ratio comparing carriers to matched controls was estimated from the 2×2 contingency table using Fisher’s exact test, which also provided the corresponding 95% confidence intervals (fisher.test, base R). Incidence rate ratios (IRR) for HA20-related condition burden were estimated using negative binomial regression (‘MASS’ package in R), with individual EHR observation period (derived from the AoU observation period table) included as an offset to account for differences in available follow-up time between cases and controls.

For comparative clinical phenotype analysis of the Pitt cohort, odds ratios and 95% confidence intervals were estimated from 2×2 contingency tables, p-values were calculated using Fisher's exact test, and p-values were adjusted for multiple comparisons using the Benjamini-Hochberg correction.

*Phenome-wide Association Studies*

We performed phenome-wide association studies (PheWAS) for individuals carrying *TNFAIP3* variants of interest (pLOF, HPPM) in the AoU and UKBB databases. ICD-9 and ICD-10 diagnosis codes were mapped to phecodes using established mappings (Phecode v1.2). Phecodes with ≥20 cases in either biobank were retained for PheWAS analysis (1,817 phecodes) ^26^^,^^27^. Diagnosis codes in AoU include both inpatient and outpatient encounters, whereas UKBB contains only inpatient diagnosis data. Therefore, we defined cases as individuals with at least two occurrences of a phecode in AoU and at least one occurrence in UKBB. Phecodes with ≥20 cases in either biobank were tested for association with qualifying TNFAIP3 variants (pLOF and functionally validated hypomorphic missense [fHPPM] variants, analyzed separately and combined) in AoU and UKBB using logistic regression with Firth correction implemented in REGENIE. Models were adjusted for sex, age, and the first 10 genetic ancestry principal components, with the leave-one-chromosome-out (LOCO) genetic predictor from REGENIE step 1 included as an offset. Within each biobank, single-variant association statistics were generated; these summary statistics, together with cohort-specific linkage disequilibrium information, were combined in a fixed-effects gene-based meta-analysis using REMETA. Reported odds ratios represent the burden-test association between qualifying *TNFAIP3* variants and each phecode. Phenome-wide statistical significance was defined using a Bonferroni-corrected threshold of p < 2.75 x 10^-5^ (0.05/1,817). In a secondary analysis, we performed a meta-PheWAS on hypomorphic missense *TNFAIP3* variants, originally identified in the Pittsburgh cohort, that were also present in AoU and UKBB. We manually annotated phecodes to classify them as inflammatory vs. non-inflammatory conditions (**Supplementary Data 4**). To assess enrichment of conditions of interest among the meta-PheWAS results, we categorized 1,876 phecodes as inflammatory, immunodeficient, or hematologic malignancy then adapted the Kolmogorov–Smirnov-like statistic from the Gene Set Enrichment Analysis (GSEA) framework, implemented using the R package ‘fgsea’.

*Prevalence Analysis of All Human Haploinsufficiency Genes*

To estimate prevalence of deleterious variants, gnomAD v4.1 exome sites data (gs://gcp-public-data--gnomad/release/4.1/ht/exomes/gnomad.exomes.v4.1.allele_number_all_sites.ht) and constraint metrics tables (gs://gcp-public-data--gnomad/release/4.1/constraint/gnomad.v4.1.constraint_metrics.ht) were retrieved. Only variants in coding regions with canonical transcript consequences were retained. For each variant, the associated gene symbols, Ensembl gene IDs, transcript IDs, and consequence terms from Variant Effect Predictor (VEP) annotations were extracted. Only variants associated with Ensembl transcripts (ENST prefix) were retained for analysis. Two classes of variants were defined based on the following criteria:

LOF variants:

- High confidence predicted loss-of-function (pLoF): frameshift, nonsense, or splice with a SpliceAI > 0.9

LOF + Damaging missense variants:

- High-confidence pLoF variants, OR
- Missense variants with CADD-Phred score > 30

Additionally, both classifications required absence of homozygotes and MAF ≤ 0.01% for the variants. For each gene, the prevalence of deleterious variants was estimated by summing the total number of heterozygous carriers across all deleterious variants and dividing by the median number of individuals with high-quality genotype calls (allele number/2) and by multiplying by 10,000.

*Cell Culture and Functional Assays*
A20‑deficient HEK293 cells were generated by co-transfection of A20 CRISPR/Cas9 KO plasmids (sc-400447) and the corresponding HDR donor plasmid using Lipofectamine 2000, followed by puromycin selection (1 µg/mL, ~4 days) and single-cell sorting of RFP-positive cells by flow cytometry. Clones were identified by PCR for HDR insertion and loss of A20 protein expression was confirmed by immunoblotting. Cells were maintained in DMEM with GlutaMAX and standard supplements. Cells were transiently transfected using FuGENE HD (Promega). For NF‑κB luciferase assays, A20‑deficient cells were co‑transfected with NF‑κB reporter plasmids (Cignal) and GFP‑tagged wild‑type or mutant A20 (GenScript, modified from Addgene Plasmid #22141). Transfection efficiency was verified at 24 hours via GFP fluorescence under an EVOS microscope. After 48 hours, cells were stimulated with TNF-α (20 ng/ml) or vehicle for 5 hours before luciferase activity was measured using the Dual-Luciferase Reporter Assay (Promega). Firefly luciferase (relative light units, RLU) was normalized to Renilla luciferase (RLU) and expressed as fold induction versus wild‑type A20. Peripheral blood mononuclear cells (PBMCs) from HA20 patients and healthy volunteers were isolated via gradient centrifugation (Sepmate) and cryopreserved. PBMCs were lysed in RIPA buffer with protease/phosphatase inhibitors (ThermoFisher). Lysates were run on Mini-Protean TGX gels (BioRad), transferred to nitrocellulose membranes, blocked with 5% NFDM, and incubated overnight with primary A20 N-terminus antibody (CST). Membranes were then washed and incubated with HRP-conjugated secondary antibodies for 1 h. After washing, bands were visualized via chemiluminescence (SuperSignal West Pico) and normalized to β-actin. PBMCs were also thawed and cultured in standard complete medium (RPMI supplemented with 10% fetal bovine serum) in 96-well U-bottom plates at 5×10⁵ cells per well in a final volume of 110 µL and stimulated with lipopolysaccharide (LPS; 1 µg/mL) for 5 hours. Supernatants were then collected and diluted prior to analysis. IL-1β concentration was measured by ELISA (R&D Systems, #DY201) according to the manufacturer's instructions.

*Statistical Analysis*

Probability loss of function intolerance (pLI) and predicted Loss-of-Function Observed/Expected Upper bound Fraction (pLOEUF) scores were calculated as previously described^28^^,^^29^. Prevalence estimates were reported with 95% confidence intervals calculated using the Wilson-Brown method (GraphPad Prism v10). Odds ratios for condition counts (manual comparative clinical phenotyping) were reported with 95% confidence intervals calculated by the conditional maximum likelihood method based on the noncentral hypergeometric distribution. For domain analysis, statistical significance was assessed using Fisher's exact test on the 2×2 contingency table of variants within vs. outside the domain by domain vs. non-domain positions, with Benjamini-Hochberg correction across domains (GraphPad Prism v10, RStudio). Group comparisons for counts of conditions and incidence rate ratios (IRRs) used negative binomial regression from the R package ‘MASS’, with individual EHR observation period included as an offset. For comparative phenotype analysis, odds ratios and generation of Forest plots used the base R function fisher.test and R package ‘metafor’, respectively. For biomarker profiling of the Pittsburgh cohort, we tested the assumptions of normality and lognormality using four methods in GraphPad Prism (D'Agostino-Pearson, Anderson-Darling, Shapiro-Wilk, and Kolmogorov-Smirnov). Where normality and lognormality were violated consistently across all four tests, the Kruskal-Wallis test with Dunn's multiple-comparison correction was used to compute p-values among the healthy, missense, and LOF groups. Where normality or lognormality were preserved, Brown-Forsythe and Welch ANOVA with Dunnett's multiple-comparison correction was used to compute p-values among the same groups.
